# How are children’s perceptions of the home environment associated with a general psychopathology factor across childhood?

**DOI:** 10.1101/2024.08.06.24311560

**Authors:** Jack K. Nejand, Margherita Malanchini, Ivan Voronin, Thalia C. Eley, Kaili Rimfeld

## Abstract

**Background:** Comorbidity and heterogeneity in psychiatric disorders may stem from a general psychopathology (p) factor influenced by both genetic and environmental factors. Although the relative contributions of these influences on psychopathology are established, the longitudinal associations between p-factor and specific environmental exposures across development are not well understood. Using a longitudinal genetically informative design, this study investigates the association between the home environment and p-factor across childhood.

**Methods:** Data were obtained from the Twins Early Development Study (TEDS). Cross-lagged panel analyses were conducted separately to ascertain the direction of associations between parent-rated p, self-rated p, and self-rated home environment (chaos at home and parental discipline) at ages 9, 12, and 16 (N=6,213). Biometric autoregressive cross-lagged twin models were used to assess the aetiology of these associations, and MZ differences analyses were used to control for familial effects.

**Results:** Both latent factors were stable over time, although twin-rated p-factor (*r* = 0.44-0.40) was more variable than parent-rated p-factor (*r* = 0.72-0.63). ‘Home environment’ was more variable than p-factor uniformly. Small, significant bi-directional associations were found between p-factor and home environment, with stronger cross-lagged paths from p-factor to home environment than vice versa. These longitudinal associations persisted over time, though attenuated for parent-rated p-factor. Genetic analyses revealed that bi-directional cross-lagged paths were largely explained by shared environmental factors, with a smaller proportion explained by genetic factors. This pattern of results was confirmed in MZ differences analyses.

**Conclusions:** Our findings suggest a dynamic and bidirectional relationship between p-factor and the home environment across development, predominantly influenced by shared environmental factors. Changes in one can influence the other, highlighting the complexity of psychopathology’s environmental influences. This underscores the need for further investigation into gene-environment interplay to inform approaches to psychopathology prevention and intervention.

**Key points and relevance:**

1. The relationship between p-factor and the home environment is dynamic and bidirectional, indicating that changes in one can influence the other across different developmental stages. However, the effect sizes of these relationships were modest.
2. Shared environmental factors played a major role in driving cross-lagged associations between p-factor and the home environment, with some genetic contribution, suggesting that the family environment can significantly shape this relationship.
3. These findings necessitate deeper investigations into gene-environment interplay in shaping psychopathology. A better understanding of these dynamics could inform effective prevention and intervention strategies for developmental psychopathology.

## Introduction

The prevalence of psychopathology amongst young people continues to rise, with 1-in-6 UK-based 6–19-year-olds experiencing identifiable mental health needs in 2021, up from 1-in-9 in 2017 (NHS Digital, 2021). Furthermore, a 2017 report identified 5% of British under-20s met criteria for multiple mental health disorders (NHS Digital, 2017), with rates likely to be much higher today. The overlap between disorders in adults roughly conforms to a ‘rule of 50%’, meaning that half of those with one disorder are likely to meet diagnostic criteria for a second disorder, and half of this group will meet diagnostic criteria for a third and so on (Caspi et al., 2014).

Recent research suggests a single component may explain such high rates of comorbidity, labelled a general psychopathology factor or “p-factor” (Lahey et al., 2012; Caspi et al., 2014; Caspi & Moffitt, 2018). The first principal component of mental health symptoms, indicative of a p-factor, accounts for a significant proportion of variance in psychiatric disorders - ranging from 40% to 50% across different data types (Allegrini et al., 2019). Higher p-factor scores are associated with higher rates of psychiatric symptoms, and thus associated life impairment, such as suicide attempts, reliance on welfare and violence convictions (Caspi et al., 2014). Higher p-factor is also linked to lower cognitive functioning longitudinally (Von Stumm, Malanchini & Fisher, 2023).

The consistent heritability of and high genetic correlations between all psychiatric disorders supports the notion that the heritability of psychiatric disorders is largely explained by a single shared genetic factor (Kendler et al., 2011; Smoller et al., 2013). Twin studies report that genetic factors explain 20-80% of p-factor longitudinally (Selzam et al., 2018), and genetic factors also account for the relative stability across development (Allegrini et al., 2019). Genomic studies show consistent evidence of heritability for p-factor (Selzam et al., 2018; Grotzinger et al., 2022; Keser et al., 2023). However, these leave a moderate proportion of the individual differences in p-factor attributable to environmental influences (e.g., 20-52%; Allegrini et al., 2019).

The home environment provides a meaningful context whereby the environmental impact on development can be directly observed. ‘Household chaos’, defined by excessive noise, overcrowding, and an absence of routine or structure (Jaffee, Hanscombe, Haworth, Davis & Plomin, 2012), has often been associated with psychopathological outcomes. Children growing up in chaotic households have been found more likely to exhibit less behavioural control (Vrijhof, van der Voort, van Ijzendoorn & Euser, 2018), more disruptive behaviour (Jaffee et al., 2012), higher rates of depression (Tucker, Sharp, van Gundy & Rebellon, 2018) and fewer social skills (Bobbitt & Gerschoff, 2016).

Less is known about the differences between siblings’ perceptions of the home environment, how this varies over time, and thus affects later psychopathological outcomes. However, their perceptions tend to better predict their self-reported psychopathology compared with parental reports (Human, Dirks, DeLongis & Chen, 2016). Parenting style is also associated with outcomes similar to household chaos, maternal negativity, and harshness, which are associated with increased internalising and externalising problems in children (Chen, Deater-Deckard & Bell, 2014). However, there is limited research on the direction of associations between home environment and psychopathology, especially using genetically informative research designs.

Understanding the bi-directional relationship between the home environment and psychopathology is crucial; it allows us to discern whether environmental factors merely reflect underlying genetic predispositions or actively shape developmental outcomes. The concept of heritability in environmental measures, such as ‘household chaos’, suggests that children’s experiences of their environment are not independent of their genetic predispositions (Hanscombe, Haworth, Davis, Jaffee & Plomin, 2011). This introduces the notion of gene-environment correlation, where genetic factors co-occur with certain environmental exposures, in turn affecting psychopathological development (Agnew-Blais et al., 2022; Knafo & Jaffee, 2013; Knopik et al., 2017). For example, children’s behaviours may evoke certain parenting styles which are thus heritable (Eley, Napolitano, Lau & Gregory, 2010; Avinun & Knafo, 2014), pointing to a possible genetic and bi-directional relationship between the home environment and psychopathology, or p-factor.

Our understanding of how genetic and environmental factors contribute to psychopathology is still developing. Here, we study the relationship between the home environment, measured as parental discipline and chaos in the household, and psychopathology, aiming to clarify how they may influence each other. We investigate these associations over time, offering new insights into the role of the home environment in shaping and being shaped by psychopathology. The moderate heritability of p-factor and perceptions of the home environment highlights the importance of using genetically informative designs to better understand their relationship. Doing so longitudinally also allows us to study the extent and direction of associations between these constructs, overcoming the limitations of previous cross-sectional studies. Genetic cross-lagged models were also calculated to estimate the respective variance explained by additive genetic (A), shared environmental (C), and non-shared environmental (E) effects. Finally, an MZ differences design was used to control both A and C effects. The present study was registered on the Open Science Framework (https://osf.io/2xgtd/) with the following aims:

1. To explore the association between perceptions of the home environment and p-factor, longitudinally from age 9-16, and the aetiology of these associations;
2. To test the stability of p-factor and perceptions of home environment across childhood using both parent-and child-report;
3. To identify how these associations are affected when controlling for genetic and shared environmental factors using an autoregressive cross-lagged twin model and MZ differences design.

## Methods

### Ethical Considerations

Written informed consent was obtained from parents before data collection and from TEDS participants themselves past the age of 18. King’s College London’s Ethics Committee approved the project for the Institute of Psychiatry, Psychology and Neuroscience PNM/09/10–104.

### Participants

The present study utilises data from the Twins Early Development Study (TEDS), a longitudinal twin study that recruited over 16,000 twin pairs born in England and Wales between 1994 and 1996. Despite some attrition, more than 10,000 twin pairs remain actively involved in the study. This cohort remains representative of the wider English and Welsh population in terms of ethnic and socioeconomic factors across waves for their birth cohort (Lockhart et al., 2023). Rich behavioural data has been collected from the twins, their parents and teachers over three decades.

The present study used all available longitudinal data from 18,548 participants (9,274 twin pairs), with up to 6,212 pairs completing any one measure. The missing data was especially evident from age 16 data collection, as data was collected only from 2 out of 4 birth cohorts during this data collection wave. Full information maximum likelihood was used to account for missing data. Using both members of the twin pair in any analysis artificially inflates the sample size, so a random member of each pair was selected for the phenotypic analyses. This sample was 50.9% female, and 34.4% were from monozygotic pairs. Analyses were conducted on measures collected at ages 9 (*M* = 9.02, SD = 0.29), 12 (*M* = 11.31, SD = 0.72) and 16 (*M* = 16.32, SD = 0.68) from twins themselves (p-factor and home environment measures) and from their parents (p-factor measures only). These ages were chosen to capture the transition from middle childhood into adolescence, and the same home environment measures were available at each, enabling longitudinal analyses.

### Measures

The present study utilises measures to reflect p-factor described in a recent TEDS study (Allegrini et al., 2019). This approach is hypothesis-free to capture as many relevant domains as possible. It includes measures not always used in the prior p-factor literature, such as those for developmental disorders. Table 1P in Supplementary Material outlines the measures used to capture p-factor and home environment composites. More information about the measures used can be found in the TEDS Data Dictionary (https://www.teds.ac.uk/datadictionary/home.htm). All p-factor measures are well-validated and have previously demonstrated sound psychometric properties, with moderate-to-high levels of internal consistency (all Cronbach’s alpha > 0.6). Different measures were used at different waves to capture parallel constructs at each wave whilst maintaining developmental appropriateness. The home environment measures (CHAOS and parental discipline, Table 1P in Supplementary Material) were chosen as they have been consistently administered at each selected wave.

### Statistical analysis

#### Descriptive statistics and sex differences between measures

Means and standard deviations were calculated for each phenotypic variable (see Supplementary Table S1). Sex differences between variables were also calculated using ANOVA.

#### Extracting p-factor and home environment composites via Factor Analyses

To model p-factor, we conducted confirmatory factor analyses (CFA) using the lavaan package with full information maximum likelihood to account for missing data. All mental health measures showed high loadings on the first factor, indicating a coherent structure for p-factor, as previously demonstrated by Allegrini et al. (2019). The primary goal of using CFA was not to identify the best-fitting model for developmental psychopathology but rather to use it as a data reduction technique to generate a general index of psychopathology.

We had only two variables for the home environment, so CFA was not feasible. Instead, we used principal component analysis (PCA), which indicated that both components had equal loading onto the first principal component. Therefore, the mean of the two variables was used as the home environment index at each age.

#### Measuring the association between the p-factor and home environment

Correlations between measures were calculated to observe levels of association. Phenotypic, genetic and MZ differences analyses were conducted using cross-lagged panel modelling. Cross-lagged effects represent the relationship between two variables when their stability and associations over time are controlled (Cole & Maxwell, 2003; Malanchini et al., 2017). This method is, therefore, appropriate for estimating bidirectional, longitudinal relationships, in this case, between p-factorand home environment after accounting for each measure’s stability and the cross-sectional relationship at previous time points. Models were estimated for both parent- and twin-rated p-factor at ages 9, 12 and 16. Informant ratings were kept separate given that very few associations between parent- and child-rated p-factor measures indicated commonality across raters (*M*=0.17 range = −0.04, 0.52]). This corroborates the frequently reported discrepancy between parent- and child ratings of psychopathology (e.g., De Los Reyes et al., 2011; 2012). Figure 1 shows the basic structure of the model for the present study. We used bootstrapped 95% confidence intervals to interpret the strengths of the cross-lagged, cross-sectional, and stability paths.

**Figure 1.**
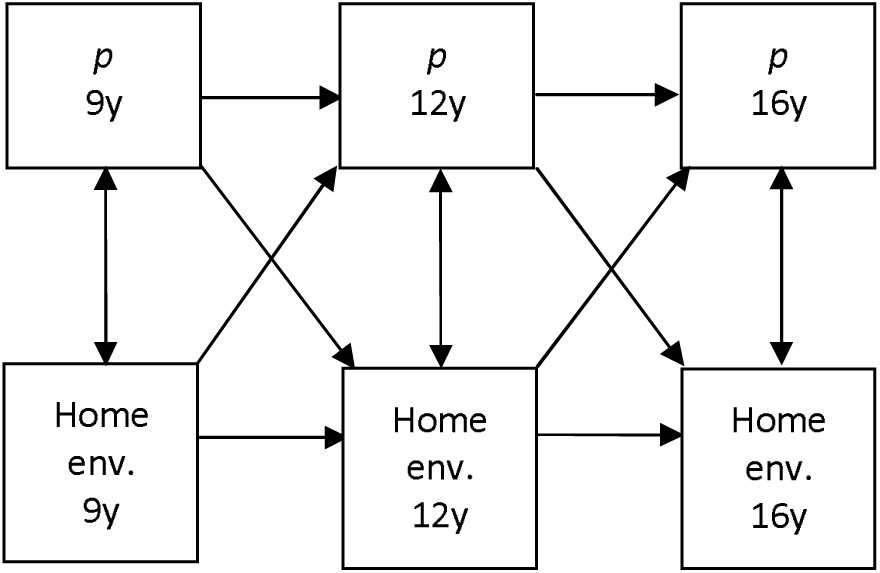
A cross-lagged panel model of p-factor and home environment across three time points (age 9, 12, 16)

#### Examining the aetiology of the longitudinal associations between p-factor and home environment

We used the twin method to examine the aetiology of p-factor and home environment and the covariation between them. Twin methods capitalise on the genetic relatedness between monozygotic or identical (MZ) and dizygotic or non-identical (DZ) twins. MZ twins share 100% of their genes, while DZ twins share, on average, 50% of their segregating genes. However, both sets of twins share their rearing environment. By comparing the difference between MZ and DZ correlations, it is possible to estimate the relative contribution of genetic, shared environmental or non-shared environmental effects to individual differences in the trait of interest. Heritability (A), the proportion of variance explained by the genetic factors, can be calculated by doubling the difference between MZ and DZ twins. Shared environmental factors (C), which make children growing up in the same family similar beyond those explained by genetic factors, can be calculated by deducting heritability from the MZ correlation. The rest of the variance is explained by non-shared environmental factors (E), environmental factors that do not contribute to similarities between twins growing up in the same family, which can be calculated by deducting MZ correlations from unity, which also includes measurement error. The ACE estimates can be calculated more accurately, including 95% confidence intervals using structural equation modelling (Knopik et al., 2017; Martin & Eaves, 1977; Rijsdijk & Sham, 2002). The present study used an OpenMx package in R for all twin analyses (Boker et al., 2011).

Univariate twin analyses can be extended to bivariate genetic analyses to study the aetiology of covariance between traits. The covariance between traits can be decomposed to additive genetic (A), shared environmental (C) and non-shared environmental (E) components by comparing the cross-twin cross-trait correlations between MZ and DZ twin pairs (Rijsdijk & Sham, 2002). This method was used to identify the aetiology of each cross-lagged association between p-factor and home environment. This analysis was not included in the pre-registration but was later determined to be an important contribution.

The cross-lagged ACE model, the TwinAnalysis package for R (https://github.com/IvanVoronin/TwinAnalysis), is an implementation of a multivariate twin model. It uses the cross-trait cross-twin correlations in MZ and DZ twin pairs to estimate the cross-lagged relationships between the additive genetic, shared environmental and non-shared environmental factors underlying the relationships between variables. These estimates are then used to calculate the proportion of phenotypic variance, explained by A, C and E. Such an approach, detailed by Malanchini et al. (2017) and utilised in subsequent studies (e.g., McAdams et al., 2020), offers causal inferences by integrating all path analyses within a single model, facilitating direct comparisons of cross-trait influences.

#### MZ differences design

The MZ differences design was also used to confirm the robustness of our findings by isolating non-shared environmental factors while controlling for genetic and shared environmental influences, allowing us to assess whether the results align with those from the cross-lagged model. Given that MZ twins share 100% of their genetics and shared environment, any differences between them are attributable to non-shared environmental influences (Liang & Eley, 2005; Richie, Bates & Plomin, 2015).

## Results

### Phenotypic analyses

#### Descriptive statistics

Means and standard deviations were calculated for measures for the whole sample, males and females, separately; these are presented in Supplementary Table S1. Analyses of variance (ANOVA) were used to test the significance of these group differences. Significant sex differences emerged for all variables, except home environment variables at age 16 and MFQ at age 12. The effect sizes of these sex differences were modest, with sex explaining, on average, around 1% of the variance, except 10% of the variance for SDQ emotional problems at age 16, which is in line with prior research (e.g., Eme, 1979; Rutter, 1988). Measures were subsequently corrected via multiple regressions for sex and age. These age and sex-corrected standardised residuals were used in all downstream analyses.

#### Correlations

As expected, the psychopathology measures were moderately correlated amongst their respective raters across ages. Figures S1-3 present the correlation heatmaps for all adjusted phenotypic variables at waves 9, 12 and 16, respectively, including correlations between phenotypic measures for single informants and child-rated home environment measures.

Figure S4 shows the correlations between the p-factor and home environment composites at each age and by each rater. Parent-rated p-factors correlated positively over time (*r* = 0.53-0.71), whereas twin-rated p-factor correlations ranged from weak to moderate over time (*r* = 0.21-0.46). p-factor correlations between raters were also moderate (*r* = 0.23-0.46). Twin-rated home environment factors were also moderately positively correlated with each other (*r* = 0.25-0.44) and also correlated with parent-rated p-factors (*r* = 0.18-0.30, *M* = 0.18) and twin-rated p-factors (*r* = 0.14-0.40).

#### Computing latent factors of mental health (p-factor)

We calculated a p-factor, i.e., a general factor for psychopathology for each age group, using confirmatory factor analyses (CFA). Here, we were not interested in the structure of mental health problems across childhood but used CFA as a data reduction technique to derive p-factors across raters to index mental health problems at every age. All psychopathology measures loaded onto the child-rated p-factor at all ages. The same applies to parent-rated p-factor except for the AQ Attention to Detail scale at age 16. The factor loadings and model fit statistics are presented in Supplementary Table S2.

#### Cross-lagged analyses

Parent-rated p-factor model. All autoregressive, cross-sectional, and cross-lagged paths were statistically significant, as presented in Figure 2. Both latent factors demonstrated stability over time with less variability in p-factor compared to the home environment. After controlling for the stability of p-factor and home environment and their cross-sectional correlations, the cross-lagged paths were also significant. Although the effect sizes were small, on average the p-factor had a larger effect on the home environment at both time points (9 to 12, β =0.18 (95%CI: 0.15-0.19); 12 to 16, β = 0.10 (95%CI: 0.07-0.12)) than the reverse (9 to 12, β = 0.04 (95%CI:0.02-0.05); 12 to 16, β = 0.08 (95%CI:0.06-0.09).

**Figure 2.**
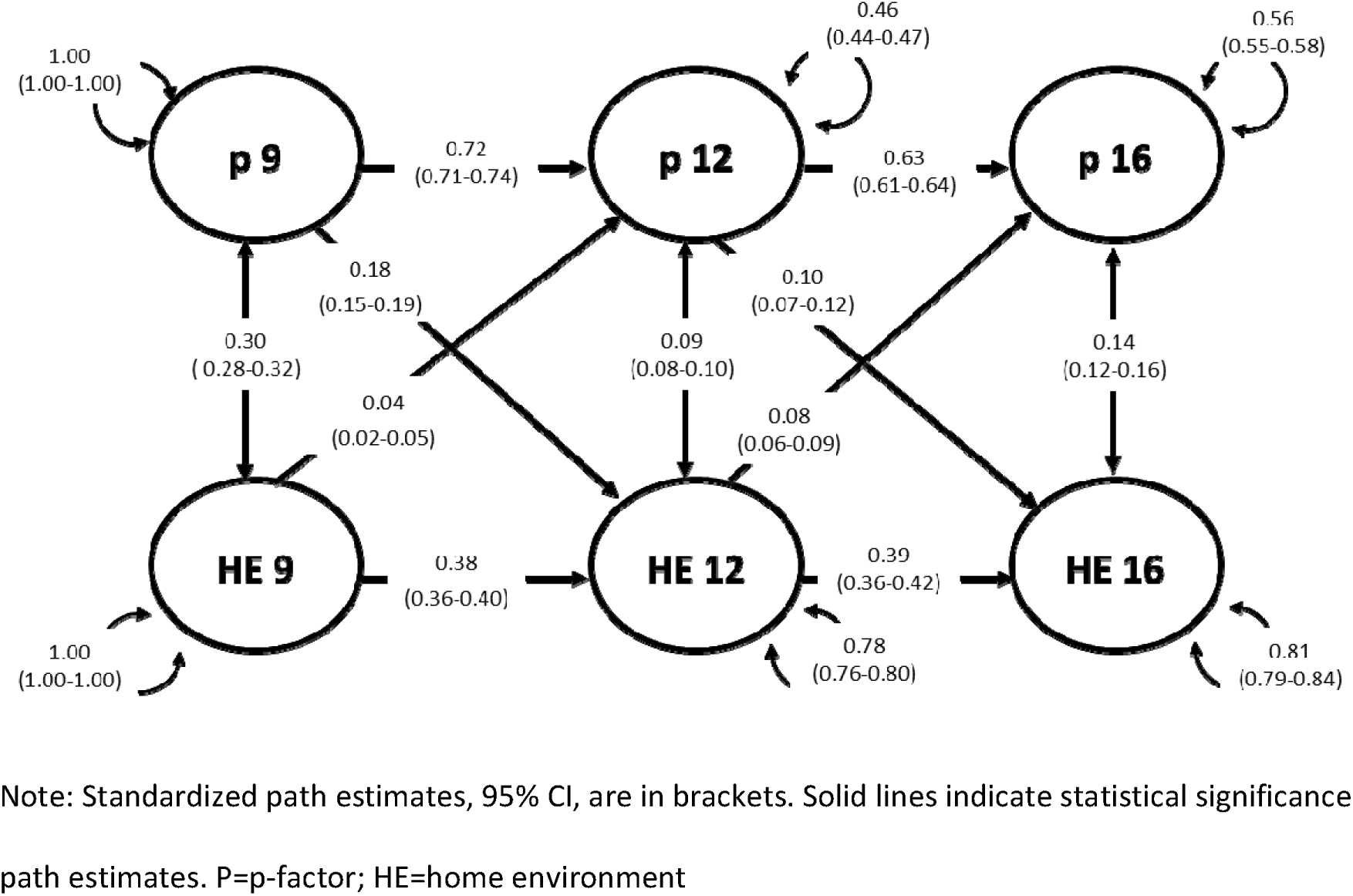
A cross-lagged panel model between the parent-rated p factor and home environment composite.

Twin-rated p-factor model. Except for the pathway home environment at 12 and p-factorat 16, all autoregressive, cross-sectional, and cross-lagged paths were again statistically significant, as illustrated in Figure 3. Both latent factors demonstrated stability over time with less variability in p-factor compared to the home environment. There was a bidirectional relationship between p-factor at 9 and HE at 12 (β = 0.15 (95%CI: 0.12-0.16)) and from HE at 9 to p-factor at 12 (β = 0.11 (95%CI: 0.09-0.13)) after controlling for stability of both constructs and their cross-sectional correlations.

**Figure 3.**
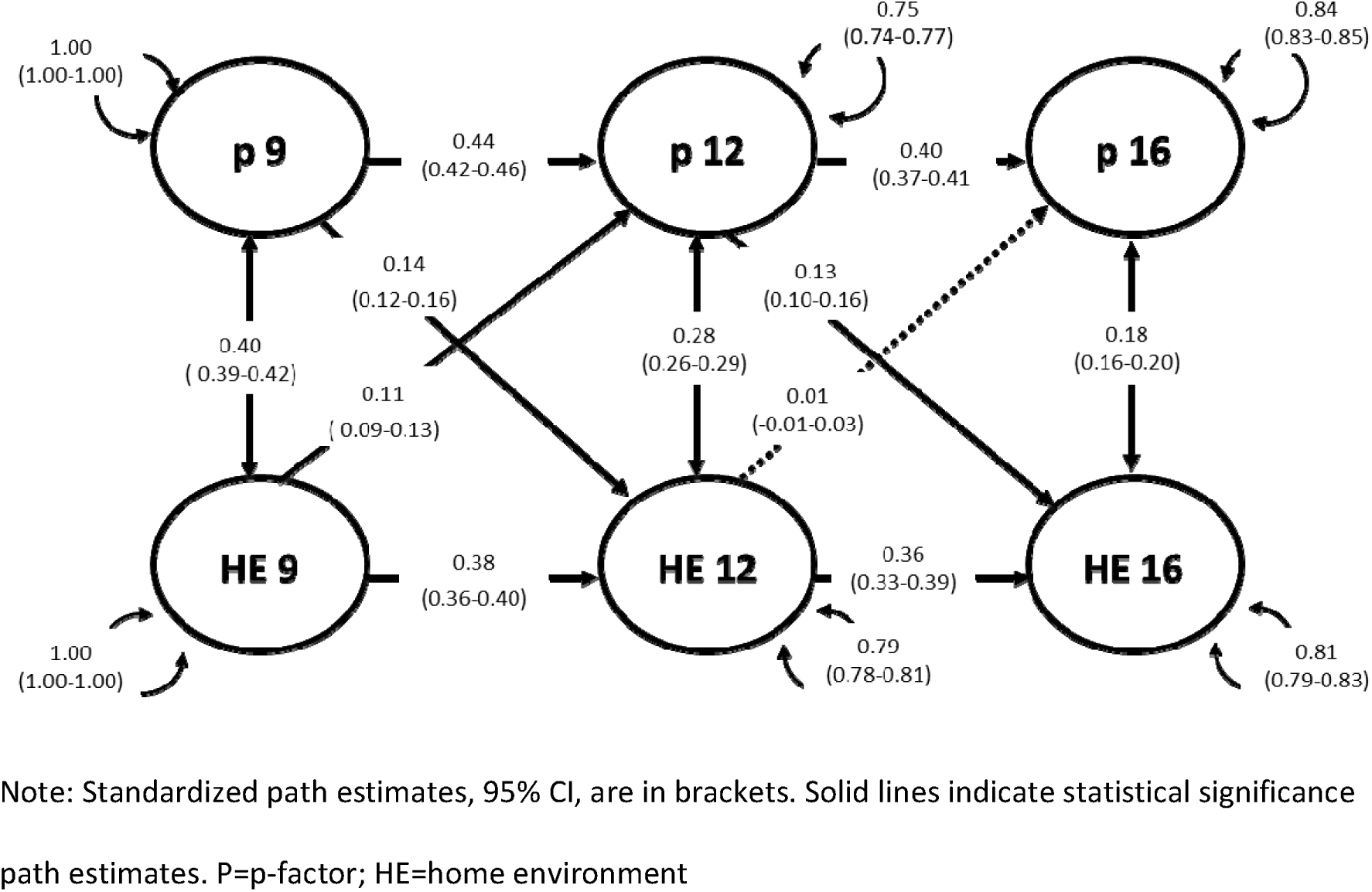
A cross-lagged panel model between the twin-rated p factor (p) and home environment composite.

However, there was only a unidirectional relationship from p-factor at 12 to the home environment at 16 (β = 0.13 (95%CI: 0.10-0.16)).

Model fit statistics for phenotypic cross-lagged models are presented in Supplementary Table S3.

### Genetic Analyses

#### ACE cross-lagged analyses

According to the fit statistics, all models provided an accurate description of the data (Supplementary Table S6).

#### Parent-rated p-factor ACE model

The heritability of parent-rated p-factor was consistently high (age 9 - h^2^ = 0.51 [0.47-0.58]; age 12 - h^2^ = 0.67 [0.63-0.71]; age 16 - h^2^ = 0.59 [0.55-0.64]), whereas the heritability of home environment was consistently lower (age 9 - h^2^ = 0.19 [0.13-0.27]; age 12 - h^2^ = 0.19 [0.12-0.24]; age 16 - h^2^ = 0.31 [0.21-0.41]). The remaining variance was accounted for by shared and non-shared environmental factors (see Supplementary Table S5).

Genetic and environmental decomposition of the developmental associations between parent-rated p-factor and home environment is presented in Figure 4. The moderate stability of p-factor was mostly explained by A (69% from age 9 to 12 and 75% from 12 to 16). Home environment stability was mostly explained by C (66% from age 9 to 12 and 62% from age 12 to 16). Cross-lagged pathways from p-factor to the home environment were also largely explained by shared environmental factors, with a smaller proportion explained by genetic factors.

**Figure 4.**
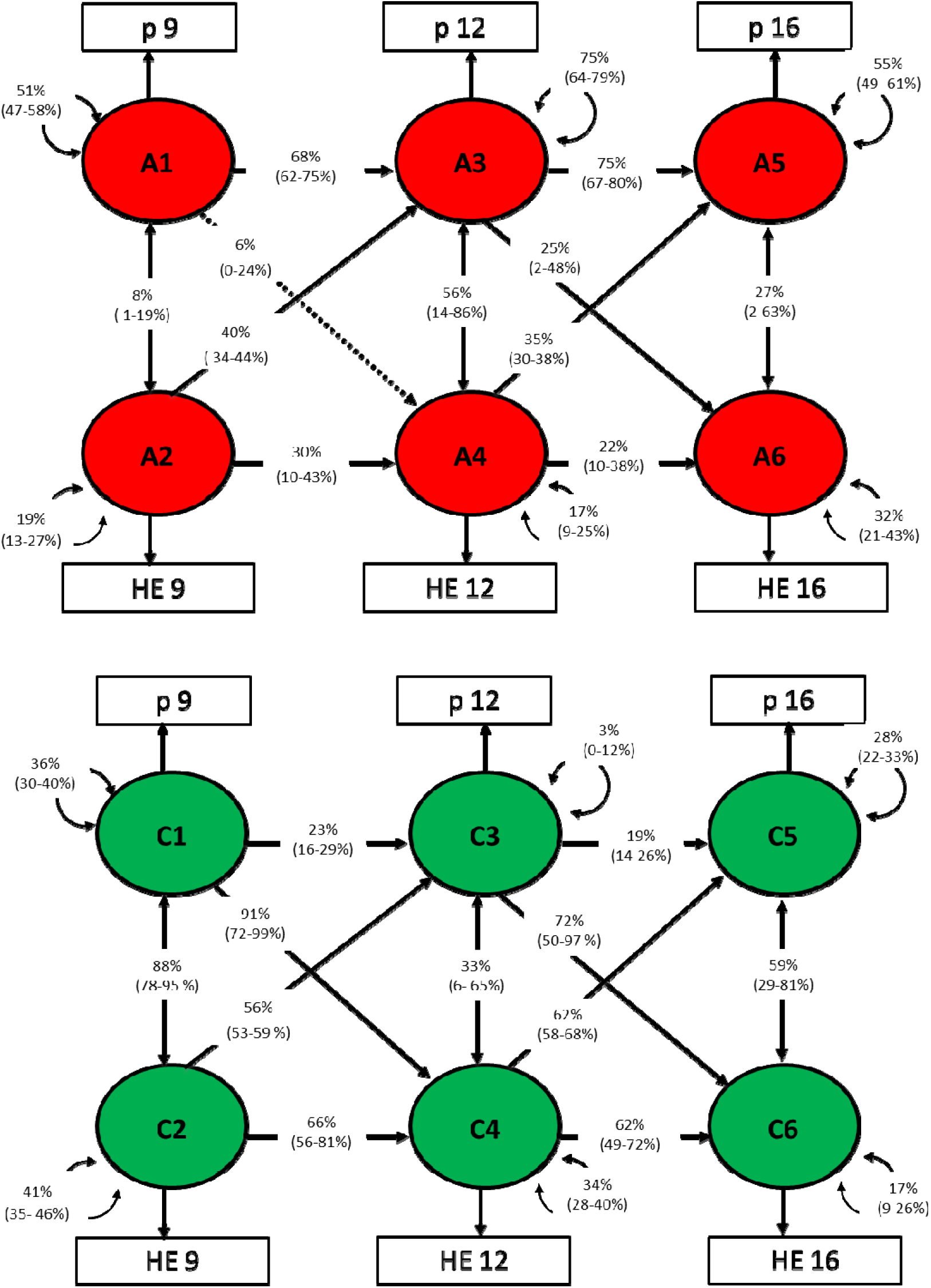
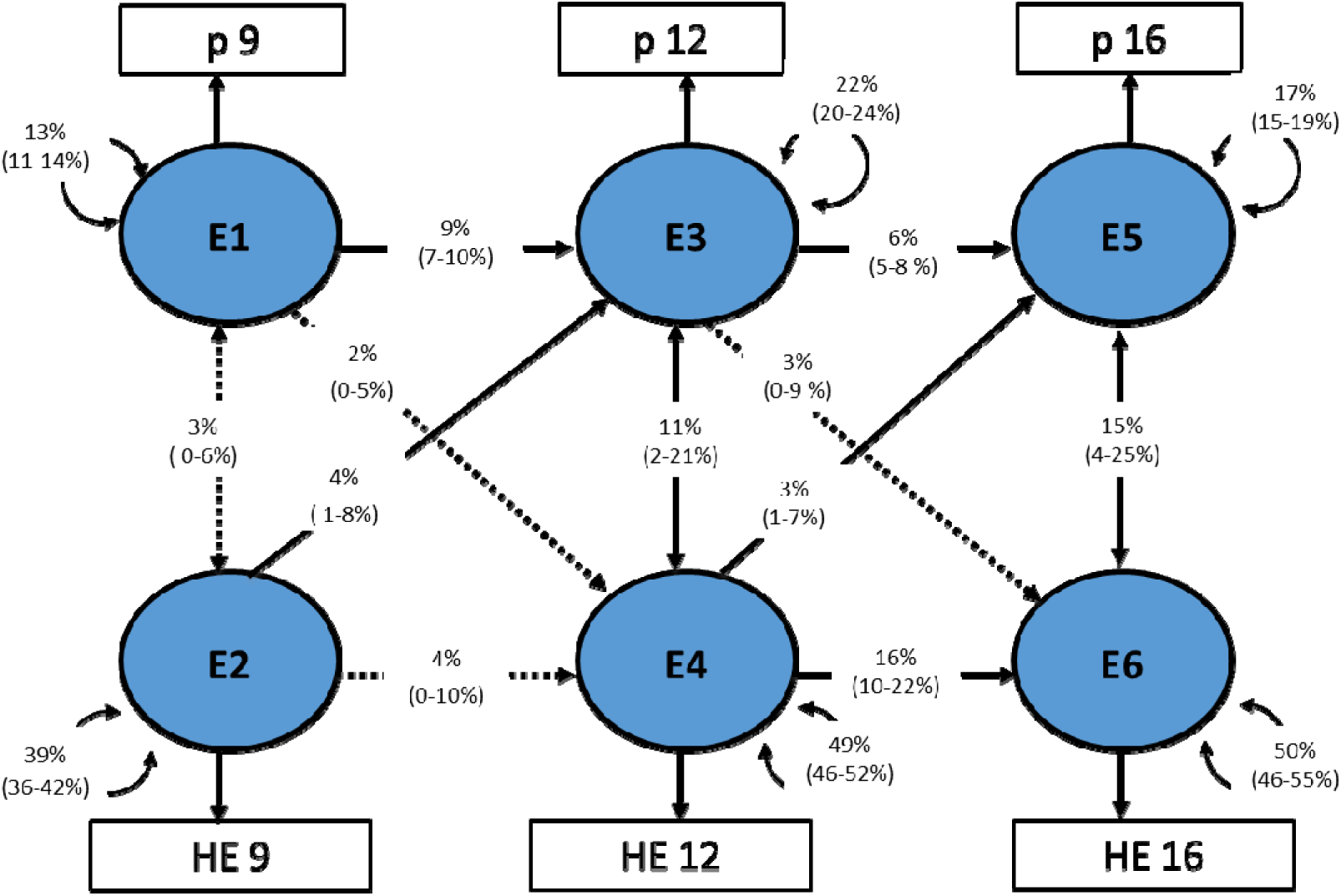
Genetic and environmental decomposition of associations between parent-rated p factor and twin-rated home environment at ages 9, 12 and 16. ‘A’ represents the proportion of variance (%) explained by additive genetic effects, ‘C’ by shared environment and ‘E’ by non-shared environment.

#### Twin-rated p-factor ACE model

The heritability of twin-rated p-factor was moderate (age 9 - h = 0.47 [0.4-0.51]; age 12 - h = 0.42 [0.37-0.47]; age 16 - h =0.46 [0.41-0.51]). The remaining variance was accounted for by shared and non-shared environmental factors (see Supplementary Table S7).

Genetic and environmental decomposition of the developmental associations between twin-rated p-factor and home environment is presented in Figure 5. The moderate stability of p-factor was mostly explained by A (77% from age 9 to 12 and 76% from 12 to 16). Home environment stability was mostly explained by C (74% from age 9 to 12 and 70% from age 12 to 16). Cross-lagged pathways from p-factor to the home environment were also largely explained by shared environmental factors, except the cross-lagged path from p-factor at 12 to the home environment at 16, which was 51% explained by genetic factors, 33% by shared environmental factors, and 16% by non-shared environmental factors.

**Figure 5.**
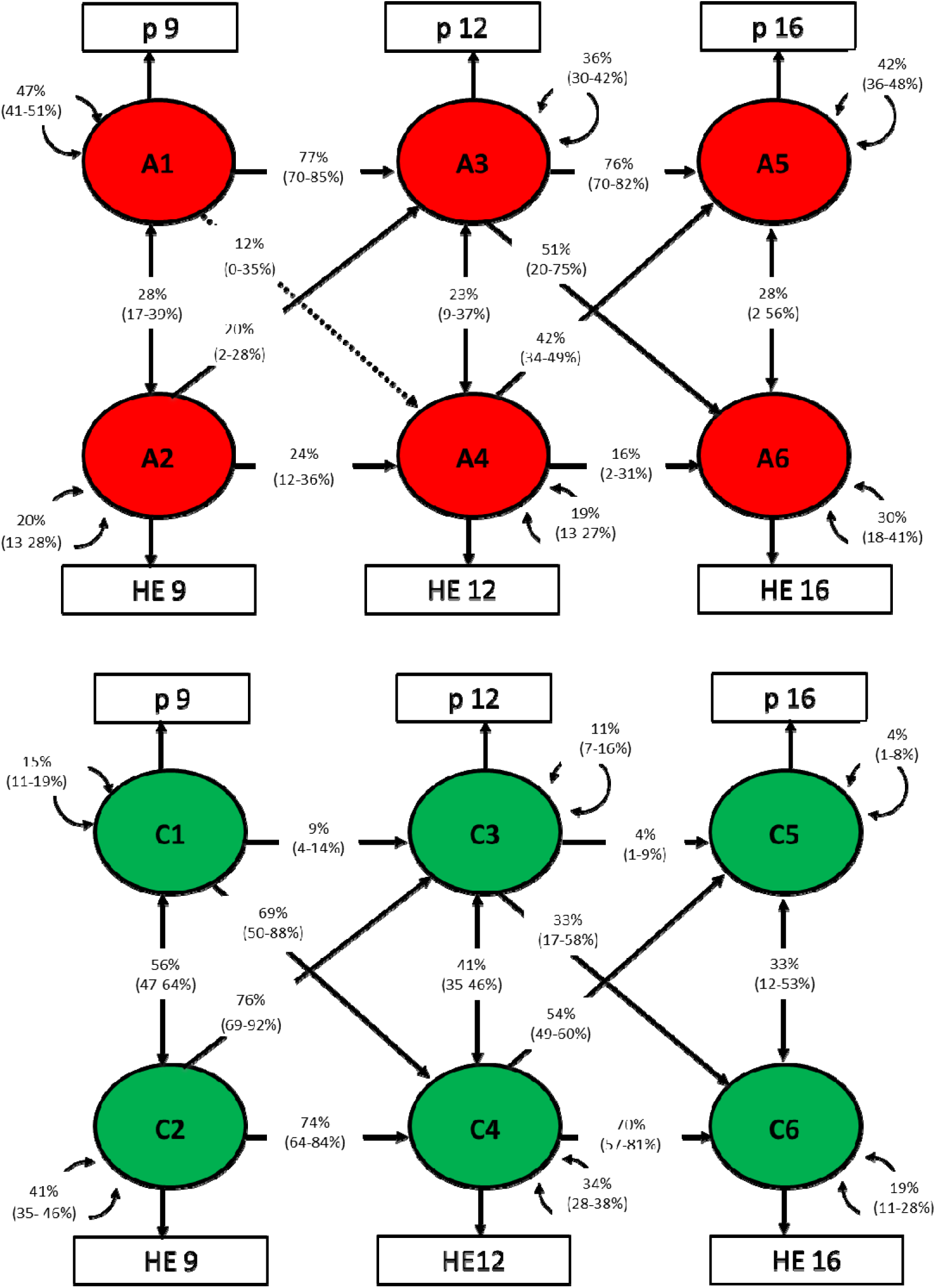
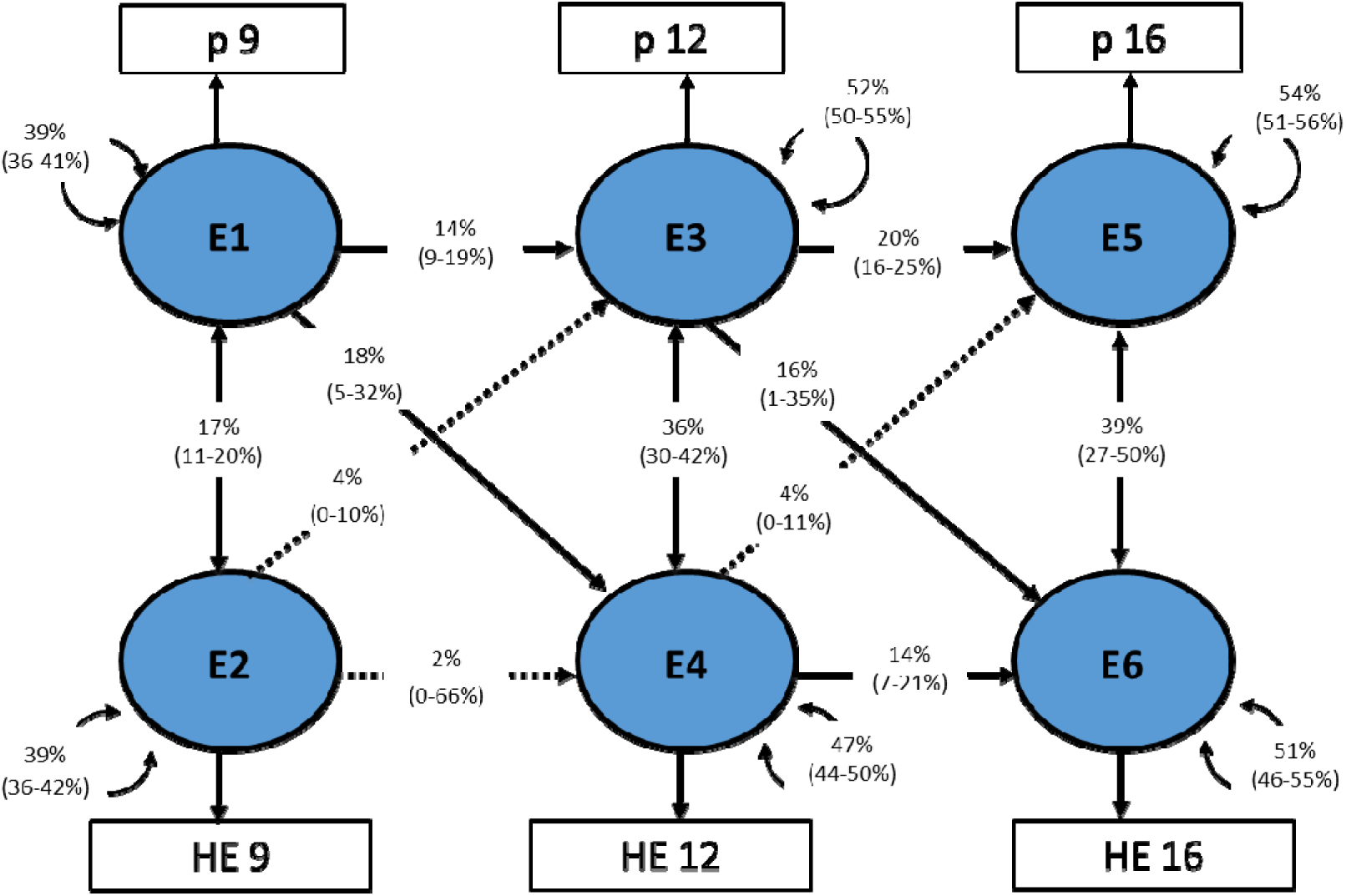
Genetic and environmental decomposition of associations between twin-rated p factor and twin-rated home environment at ages 9, 12 and 16. ‘A’ represents the proportion of variance (%) explained by additive genetic effects, ‘C’ by shared environment and ‘E’ by non-shared environment.

### MZ differences analyses

#### Parent-rated p-factor model

No significant cross-lagged paths were found between parent-rated p-factor and home environment when controlling for genetics and shared environment (see Figure S5). The p-factor remained stable over time (mean β = 0.23), whereas the home environment only demonstrated stability from age 12 to 16 (β = 0.10). Cross-sectional paths only became statistically significant at ages 12 and 16, but again were small (mean β = 0.07).

#### Twin-rated p-factor model

Except for the pathway from p-factor at 9 to the home environment at 12 (β = 0.09), no significant cross-lagged paths were found between the twin-rated p-factor and the home environment (see Figure S6). As before, p-factor remained stable over time (mean β = 0.10), whereas the home environment only demonstrated stability from age 12 to 16 (β = 0.09), but estimates were small in both cases. All cross-sectional paths were small but statistically significant (mean β = 0.17) and attenuated over time.

## Discussion

Using a cross-lagged panel model, our study explored the longitudinal associations between child-reported home environment and general psychopathology (p) factor during key developmental phases. The results illustrate those perceptions of the household environment, specifically chaos and parental discipline, are predictive of later psychopathological outcomes. Conversely, early indicators of the p-factor can forecast subsequent household conditions. Notably, the reciprocal relationships between these domains were observed independently of their concurrent intercorrelations and stability. Although the effects identified were modest, their statistical significance suggests that interventions to improve household dynamics might also positively impact psychopathological trajectories.

Our study’s robustness stems from genetically informed designs, including genetic cross-lagged models and an MZ differences design. This approach revealed that the stability of p-factor over time was mainly due to genetic influences, while the shared environmental factors mainly explained the relative stability of the home environment. In contrast, shared environmental factors predominantly shaped the bidirectional influences between these two factors. These findings were further substantiated by the MZ differences design, which controls for both genetic and shared environmental variables, demonstrating that the cross-lagged influences were not significant when these factors were controlled for.

Despite modest correlations between parent and twin ratings of p-factor, the findings were remarkably consistent across both models. Both demonstrated high heritability for p-factorand moderate stability over time, primarily driven by genetic factors. The twin-rated model showed less stability in p-factor compared to the parent-rated model. Shared environmental factors largely explained the cross-lagged pathways in both models, but genetic factors contributed more significantly to the twin-rated model, especially for the path from the p-factor at age 12 to the home environment at age 16.

The current findings reiterate the importance of multi-informant assessment. The analyses revealed that child-reported home environment at age 12 is associated with p-factor at 16 in the parent-rated model but not in the twin-rated model. The use of age-appropriate measures for young people across development, alongside valid parental measures for the same constructs remains key. Although effect sizes were small here, providing psychoeducation around the typical traits that evoke chaotic households for caregivers and environmental triggers of behaviour for children can still be a discrete intervention target. This was demonstrated by a mindfulness-based parenting programme where parental (*t* = −5.3) and child psychopathology outcomes (*t* = −4.3) and parental stress (*t* = −5.7) all improved significantly up to 8 weeks later (Bögels, Hellemans, van Deursen, Römer & van der Meulen, 2014). Moreover, the likely genetic influence found here may indicate that family-based transdiagnostic treatment can yield collective positive outcomes, perhaps by focusing on shared aetiological factors between family members. Establishing standardised assessments of the home environment and genetic traits such as caregiver psychopathology may flag appropriate families for whom this approach is most beneficial.

The general factor of psychopathology, ‘p’, represents commonalities across mental disorders, derived statistically from symptom correlations. It is a statistical construct, and there is no consensus on what p-factor really is, which limits the present study’s findings (see Lane, Steinley & Sher, 2016; Watts, Lane, Bonifay, Steinley & Meyer, 2020). More widely, a consensus on measuring p-factor is needed (Fried, Greene & Eaton, 2021; Lahey, Moore, Kaczkurkin & Zald, 2021). This may come via dimensional rather than diagnostic measures or norms weighted by their loadings onto p-factor (Pettersson et al., 2018). Treatment modality may also be discernible where individuals with broad comorbidity score highly on specific factors after isolating p-factor (Pettersson et al., 2021). The present study attempted to overcome this by replicating the hypothesis-free approach of a previous same-sample study (Allegrini et al., 2019). The primary aim was to investigate the longitudinal interplay between general psychopathology and home environment, not to verify the psychopathology’s underlying factor structure.

### Limitations and future directions

There are several limitations to consider. The primary limitation of our study lies in the diversity of measurement instruments used to assess p-factor across different assessment waves. This variation prompts questions about whether observed developmental changes are due to actual shifts in the underlying constructs or differences in measurement tools. Despite using age-appropriate and valid measures, further research is necessary to ensure these tools consistently capture the same constructs over time (Von Stumm, Malanchini & Fisher, 2023).

Although we used the same home environment measures developmentally, we were limited to twin-reported household chaos and parental discipline only, and the reliance on self-reported data for assessing the home environment could introduce biases that do not fully reflect the actual dynamics within the household. A broader measure of the home environment could also account for the proximity and severity of exposure to neighbourhood disorder, housing conditions and relationship instability, all of which are indicative of future psychopathology (Coley, Lynch & Kull, 2015). Future studies might benefit from incorporating objective measurements or observational data for a more comprehensive analysis.

Parental home environment measures were omitted here but could add to clinical understanding and provide objective and subjective information to assess the overall environment. Measures such as the Home Observation for Measurement of the Environment Scale (HOME; Caldwell & Bradley, 2003) may better operationalise this construct by including physical and relational aspects (e.g., cleanliness, safety, parental responsiveness).

Another consideration is the timing of our data collection. Given that assessments were conducted every few years, important nuances in the developmental trajectory of p-factor in relation to home environment changes might have been missed. More frequent data collection could help elucidate these dynamics more clearly.

The generalisability of our findings may be also limited due to our study’s specific demographic and geographic focus. Extending this research to a broader range of populations would help verify the universality of the observed associations.

Lastly, it is argued that when stable traits such as p-factor are modelled using cross-lagged panel modelling, cross-lagged relationships are confounded by between-person differences, meaning within-person relationships are more challenging to infer casually (Hamaker, Kuiper & Grasman, 2015). An alternative methodological approach, random-intercept cross-lagged panel modelling (RI-CLPM), may be more suitable, having already supported dimensional comorbidity in 7–12-year-olds and their families (Allegrini et al., 2021).

## Conclusion

The present study aimed to build on the emerging p-factor literature by investigating its longitudinal association with the home environment. While the study points to a significant bidirectional relationship influenced by both genetic and environmental elements, it is essential to interpret these findings within the context of current methodological limitations. The complexity and variability inherent in measuring the ‘p-factor’ and the home environment caution against drawing definitive conclusions about their developmental interplay. Perhaps more evident are age-specific processes, which create a complex interplay between genes and the environment throughout development. These preliminary insights, though valuable, underscore the need for further, more detailed research to clarify these relationships and their implications for childhood psychopathology. Operationalising the relevant indicators of p-factor and home environment will improve understanding of these processes and benefit clinical practice by informing family-based interventions. We hope this study serves as a stepping stone towards a deeper understanding rather than a conclusive guide for clinical application.

**Supporting information accompanies this manuscript.**

## Data Availability

For information on data availability, please see the TEDS data access policy. All relevant data are available from the authors according to the TEDS data access policy.

http://www.teds.ac.uk/research/collaborators-and-data/teds-data-access-policy

## Acknowledgements

We gratefully acknowledge the ongoing contribution of the participants in the Twins Early Development Study (TEDS) and their families. TEDS is supported by a programme Grant to R.P. from the UK Medical Research Council (Grant Nos. MR/V012878/1 and previously MR/M021475/1), with additional support from the US National Institutes of Health (Grant No. AG046938). The research leading to these results has also received funding from the European Research Council under the European Union’s Seventh Framework Programme (FP7/2007-2013)/ grant agreement n° 602768. Project approval was granted by the IoPPN DClinPsy programme team on 20^th^ March 2020. A Twins Early Development Study (TEDS) data request was approved on 14^th^ July 2020. This required pre-registration on the Open Science Framework (https://osf.io/2xgtd/).

## Notes

### Competing Interest Statement

The authors have declared no competing interest.

### Author Declarations

Written informed consent was obtained from parents before data collection and from TEDS participants themselves past the age of 18. King's College London's Ethics Committee approved the project for the Institute of Psychiatry, Psychology and Neuroscience PNM/09/10-104.

## References

Agnew-Blais, J. C., Wertz, J., Arseneault, L., Belsky, D. W., Danese, A., Pingault, J. B., Polanczyk, G. V., Sugden, K., Williams, B., & Moffitt, T. E. (2022). Mother’s and children’s ADHD genetic risk, household chaos and children’s ADHD symptoms: A gene–environment correlation study. Journal of Child Psychology and Psychiatry, 63(10), 1153–1163.

Allegrini, A. G., Cheesman, R., Rimfeld, K., Selzam, S., Pingault, J. B., Eley, T., & Plomin, R. (2019). The p factor: Genetic analyses support a general dimension of psychopathology in childhood and adolescence. bioRxiv, 591354.

Allegrini, A., van Beijsterveldt, T., Boomsma, D., Rimfeld, K., Pingault, J. B., Plomin, R., Bartels, M., & Nivard, M. (2021). Developmental co-occurrence of psychopathology dimensions in childhood: between and within person processes. PsyArXiv.

Avinun, R., & Knafo, A. (2014). Parenting as a reaction evoked by children’s genotype: A meta-analysis of children-as-twins studies. Personality and Social Psychology Review, 18(1), 87–102.

Bobbitt, K. C., & Gershoff, E. T. (2016). Chaotic experiences and low-income children’s social-emotional development. Children and Youth Services Review, 70, 19–29.

Bögels, S. M., Hellemans, J., van Deursen, S., Römer, M., & van der Meulen, R. (2014). Mindful parenting in mental health care: effects on parental and child psychopathology, parental stress, parenting, coparenting, and marital functioning. Mindfulness, 5(5), 536–551.

Boker, S., Neale, M., Maes, H., Wilde, M., Spiegel, M., Brick, T., Spies, J., Estabrook, R., Kenny, S., Bates, T., Mehta, P., & Fox, J. (2011). OpenMx: an open source extended structural equation modeling framework. Psychometrika, 76, 306–317.

Caldwell, B. M., & Bradley, R. H. (2003). Home Observation for Measurement of the Environment: Administration manual. Tempe, AZ: Family & Human Dynamics Research Institute, Arizona University.

Caspi, A., Houts, R. M., Belsky, D. W., Goldman-Mellor, S. J., Harrington, H., Israel, S., Meier, M. H., Ramrakha, S., Shalev, I., Poulton, R. & Moffitt, T. E. (2014). The p factor: one general psychopathology factor in the structure of psychiatric disorders?. Clinical Psychological Science, 2(2), 119–137.

Caspi, A., & Moffitt, T. E. (2018). All for one and one for all: Mental disorders in one dimension. American Journal of Psychiatry, 175(9), 831–844.

Chen, N., Deater-Deckard, K., & Bell, M. A. (2014). The role of temperament by family environment interactions in child maladjustment. Journal of Abnormal Child Psychology, 42(8), 1251–1262.

Cole, D. A., & Maxwell, S. E. (2003). Testing mediational models with longitudinal data: Questions and tips in the use of structural equation modeling. Journal of Abnormal Psychology, 112(4), 558–577

Coley, R. L., Lynch, A. D., & Kull, M. (2015). Early exposure to environmental chaos and children’s physical and mental health. Early Childhood Research Quarterly, 32, 94–104.

De Los Reyes, A., Youngstrom, E. A., Swan, A. J., Youngstrom, J. K., Feeny, N. C., & Findling, R. L. (2011). Informant discrepancies in clinical reports of youths and interviewers’ impressions of the reliability of informants. Journal of Child and Adolescent Psychopharmacology, 21 (5), 417–424.

De Los Reyes, A., Thomas, S. A., Swan, A. J., Ehrlich, K. B., Reynolds, E. K., Suarez, L., Dougherty, L. R., MacPherson, L., & Pabón, S. C. (2012). “It depends on what you mean by ‘disagree’”: Differences between parent and child perceptions of parent–child conflict. Journal of Psychopathology and Behavioral Assessment, 34 (3), 293–307.

Eley, T. C., Napolitano, M., Lau, J. Y., & Gregory, A. M. (2010). Does childhood anxiety evoke maternal control? A genetically informed study. Journal of Child Psychology and Psychiatry, 51(7), 772–779.

Eme, R. F. (1979). Sex differences in childhood psychopathology: a review. Psychological Bulletin, 86(3), 574–595.

Fried, E. I., Greene, A. L., & Eaton, N. R. (2021). The p factor is the sum of its parts, for now. World Psychiatry, 20(1), 69–70.

Grotzinger, A. D., Mallard, T. T., Akingbuwa, W. A., Ip, H. F., Adams, M. J., Lewis, C. M., … & Nivard, M. G. (2022). Genetic architecture of 11 major psychiatric disorders at biobehavioral, functional genomic and molecular genetic levels of analysis. Nature Genetics, 54(5), 548–559.

Hamaker, E. L., Kuiper, R. M., & Grasman, R. P. (2015). A critique of the cross-lagged panel model. Psychological Methods, 20(1), 102–116.

Hanscombe, K. B., Haworth, C. M., Davis, O. S., Jaffee, S. R., & Plomin, R. (2011). Chaotic homes and school achievement: A twin study. Journal of Child Psychology and Psychiatry, 52(11), 1212–1220.

Human, L. J., Dirks, M. A., DeLongis, A., & Chen, E. (2016). Congruence and incongruence in adolescents’ and parents’ perceptions of the family: Using response surface analysis to examine links with adolescents’ psychological adjustment. Journal of Youth and Adolescence,

Jaffee, S. R., Hanscombe, K. B., Haworth, C. M., Davis, O. S., & Plomin, R. (2012). Chaotic homes and children’s disruptive behavior: A longitudinal cross-lagged twin study. Psychological Science, 23(6), 643–650.

Kendler, K. S., Aggen, S. H., Knudsen, G. P., Røysamb, E., Neale, M. C., & Reichborn-Kjennerud, T. (2011). The structure of genetic and environmental risk factors for syndromal and subsyndromal common DSM-IV axis I and all axis II disorders. American Journal of Psychiatry, 168(1), 29–39.

Keser, E., Liao, W., Allegrini, A. G., Rimfeld, K., Eley, T. C., Plomin, R., & Malanchini, M. (2023). Isolating transdiagnostic effects reveals specific genetic profiles in psychiatric disorders. *medRxiv,* 2023-12.

Knafo, A., & Jaffee, S. R. (2013). Gene–environment correlation in developmental psychopathology. Development and psychopathology, 25(1), 1–6.

Knopik, V. S., Neiderhiser, J. M., DeFries, J. C., & Plomin, R. (2017). Behavioral Genetics. New York: Worth Publishers, Macmillan Learning.

Lahey, B. B., Applegate, B., Hakes, J. K., Zald, D. H., Hariri, A. R., & Rathouz, P. J. (2012). Is there a general factor of prevalent psychopathology during adulthood?. Journal of Abnormal Psychology, 121(4), 971–977.

Lahey, B. B., Moore, T. M., Kaczkurkin, A. N., & Zald, D. H. (2021). Hierarchical models of psychopathology: empirical support, implications, and remaining issues. World Psychiatry, 20(1), 57–63.

Lane, S. P., Steinley, D., & Sher, K. J. (2016). Meta-analysis of DSM alcohol use disorder criteria severities: Structural consistency is only ‘skin deep.’ Psychological Medicine, 46(8), 1769– 1784.

Liang, H., & Eley, T. C. (2005). A monozygotic twin differences study of non-shared environmental influence on adolescent depressive symptoms. Child Development, 76(6), 1247–1260.

Lockhart, C., Ahmadzadeh, Y., Breen, G., Bright, J., Bristow, S., Boyd, A., Downs, J., Hotopf, M., Palaiologou, E., Rimfeld, K., Maxwell, J., Malanchini, M., McAdams, T. A., Plomin, R., and Eley, T. C. (2023). Twins Early Development Study (TEDS): A genetically sensitive investigation of mental health outcomes in the mid-twenties. JCPP Advances, 3(2).

Malanchini, M., Wang, Z., Voronin, I., Schenker, V. J., Plomin, R., Petrill, S. A., & Kovas, Y. (2017). Reading self-perceived ability, enjoyment and achievement: A genetically informative study of their reciprocal links over time. Developmental Psychology, 53(4), 698–712.

Martin, N. G., & Eaves, L. J. (1977). The genetical analysis of covariance structure. Heredity, 38(1), 79–95.

McAdams, T. A., Rijsdijk, F. V., Zavos, H. M., & Pingault, J. B. (2021). Twins and causal inference: leveraging nature’s experiment. Cold Spring Harbor Perspectives in Medicine, 11(6), a039552.

Neale, M. C., Hunter, M. D., Pritikin, J. N., Zahery, M., Brick, T. R., Kirkpatrick, R. M., Estabrook, R., Bates, T. C., Maes, H. M., & Boker, S. M. (2016). OpenMx 2.0: Extended structural equation and statistical modeling. Psychometrika, 81(2), 535–549.

NHS Digital (2017): ‘Mental Health of Children and Young People in England, 2017’. Available at: https://digital.nhs.uk/data-and-information/publications/statistical/mental-health-of-children-and-young-people-in-england/2017/2017

NHS Digital (2021): ’Mental Health of Children and Young People in England 2021’. Available at: https://digital.nhs.uk/data-and-information/publications/statistical/mental-health-of-children-and-young-people-in-england/2021-follow-up-to-the-2017-survey

Pettersson, E., Lahey, B. B., Larsson, H., & Lichtenstein, P. (2018). Criterion validity and utility of the general factor of psychopathology in childhood: Predictive associations with independently measured severe adverse mental health outcomes in adolescence. Journal of the American Academy of Child & Adolescent Psychiatry, 57, (6), 372–383.

Pettersson, E., Larsson, H., & Lichtenstein, P. (2021). Psychometrics, interpretation and clinical implications of hierarchical models of psychopathology. World Psychiatry, 20(1), 68–69.

Rijsdijk, F. V., & Sham, P. C. (2002). Analytic approaches to twin data using structural equation models. *Briefings in bioinformatic*,s3(2), 119–133.

Ritchie, S. J., Bates, T. C., & Plomin, R. (2015). Does learning to read improve intelligence? A longitudinal multivariate analysis in identical twins from age 7 to 16. Child development, 86(1), 23–36.

Rutter, M. (1988). Epidemiological approaches to developmental psychopathology. Archives of General Psychiatry, 45(5), 486–495.

Selzam, S., Coleman, J. R. I., Caspi, A., Moffitt, T. E., & Plomin, R. (2018). A polygenic p factor for major psychiatric disorders. Translational Psychiatry, 8(205), 1–9.

Smoller, J. W., Craddock, N., Kendler, K., Lee, P. H., Neale, B. M., Numberger, J. I., & Sullivan, P. F. (2013). Identification of risk loci with shared effects on five major psychiatric disorders: A genome-wide analysis. The Lancet, 381, 1371–1379.

Tucker, C. J., Sharp, E. H., Van Gundy, K. T., & Rebellon, C. (2018). Household chaos, hostile parenting, and adolescents’ well-being two years later. Journal of Child and Family Studies, 27, 3701–3708.

Von Stumm, S., Malanchini, M., & Fisher, H. (2023). The developmental interplay between the p-factor of psychopathology and the g-factor of intelligence from age 7 through 16 years. Development and Psychopathology, 1–10.

Vrijhof, C. I., van der Voort, A., van IJzendoorn, M. H., & Euser, S. (2018). Stressful family environments and children’s behavioral control: A multimethod test and replication study with twins. Journal of Family Psychology, 32(1), 49–59.

Watts, A. L., Lane, S. P., Bonifay, W., Steinley, D., & Meyer, F. A. (2020). Building theories on top of, and not independent of, statistical models: The case of the p-factor. Psychological Inquiry, 31(4), 310–313.

