## Supplementary Materials for "How are children’s perceptions of the home environment associated with a general psychopathology factor across childhood?"

|  | Supplementary Tables  Table S1. *Descriptive statistics for phenotypic variables and ANOVA results testing for sex differences (using raw scores for phenotypic analysis sample only)* | | | | | | | | | | | | | | |
| --- | --- | --- | --- | --- | --- | --- | --- | --- | --- | --- | --- | --- | --- | --- | --- |
| **Wave age** | | **Informant** | **Composite** | **Measure** | ***n*** | | | | | **Mean (SD)** | | | ***F value*** | ***p*** | ***R^2^*** |
|  |  |  |  |  | **Overall** | | **Male** | **Female** | | **Overall** | **Male** | **Female** |  |  |  |
| **9** | | Parent | P factor | SDQ (Prosocial) | 3407 | 1787 | | | 1620 | 8.28 (1.69) | 8.60 (1.52) | 7.93 (1.79) | 139.28 | <0.01 | 0.04 |
|  |  |  |  | SDQ (Hyperactivity) | 3403 | 1786 | | | 1619 | 3.28 (2.40) | 2.85 (2.18) | 3.74 (2.53) | 32.78 | <0.01 | 0.01 |
|  |  |  |  | SDQ (Conduct) | 3405 | 1786 | | | 1619 | 1.28 (1.44) | 1.15 (1.34) | 1.43 (1.53) | 25.28 | <0.01 | 0.01 |
|  |  |  |  | SDQ (Peer Problems) | 3405 | 1787 | | | 1618 | 1.08 (1.56) | 0.96 (1.40) | 1.22 (1.72) | 20.67 | <0.01 | 0.01 |
|  |  |  |  | SDQ (Anxiety) | 3405 | 1785 | | | 1618 | 1.78 (1.94) | 1.92 (1.96) | 1.62 (1.90) | 120.40 | <0.01 | 0.03 |
|  |  |  |  | APSD (Callous-Unemotional) | 3410 | 1786 | | | 1618 | 3.39 (1.86) | 3.12 (1.81) | 3.70 (1.86) | 36.86 | <0.01 | 0.01 |
|  |  |  |  | APSD (Narcissism) | 3404 | 1783 | | | 1615 | 1.88 (2.02) | 1.73 (1.92) | 2.05 (2.11) | 52.39 | <0.01 | 0.02 |
|  |  |  |  | APSD (Impulsivity) | 3409 | 1786 | | | 1619 | 3.29 (2.08) | 2.93 (1.99) | 3.68 (2.10) | 94.86 | <0.01 | 0.03 |
|  |  |  |  | RPAQ (Proactive Aggression) | 3404 | 1785 | | | 1616 | 0.37 (0.74) | 0.30 (0.68) | 0.45 (0.79) | 73.15 | <0.01 | 0.02 |
|  |  |  |  | RPAQ (Reactive Aggression) | 3398 | 1786 | | | 1617 | 2.01 (1.39) | 1.85 (1.35) | 2.19 (1.41) | 40.07 | <0.01 | 0.01 |
|  |  |  |  | CAST (Social) | 3405 | 1788 | | | 1622 | 2.43 (1.84) | 2.15 (1.70) | 2.75 (1.94) | 84.55 | <0.01 | 0.02 |
|  |  |  |  | CAST (Non-Social) | 3401 | 1786 | | | 1618 | 2.15 (1.57) | 1.93 (1.41) | 2.39 (1.70) | 20.98 | <0.01 | 0.01 |
|  |  |  |  | CAST (Communication) | 3403 | 1787 | | | 1622 | 2.38 (2.01) | 2.18 (1.84) | 2.61 (2.16) | 116.01 | <0.01 | 0.03 |
|  |  | Twin | Home environment | Parental discipline | 3326 | 1758 | | | 1568 | 3.19 (1.59) | 3.07 (1.57) | 3.33 (1.60) | 22.20 | <0.01 | 0.01 |
|  |  |  |  | CHAOS | 3394 | 1791 | | | 1603 | 4.50 (2.34) | 4.34 (2.30) | 4.68 (2.37) | 17.83 | <0.01 | 0.01 |
|  |  |  | P factor | SDQ (Prosocial) | 3371 | 1780 | | | 1591 | 7.93 (1.85) | 8.33 (1.64) | 7.48 (1.96) | 188.33 | <0.01 | 0.05 |
|  |  |  |  | SDQ (Hyperactivity) | 3370 | 1779 | | | 1591 | 3.92 (2.26) | 3.62 (2.16) | 4.25 (2.32) | 65.97 | <0.01 | 0.02 |
|  |  |  |  | SDQ (Conduct) | 3369 | 1780 | | | 1589 | 2.21 (1.84) | 1.99 (1.72) | 2.46 (1.93) | 56.96 | <0.01 | 0.02 |
|  |  |  |  | SDQ (Peer Problems) | 3354 | 1780 | | | 1574 | 1.92 (1.75) | 1.79 (1.70) | 2.06 (1.80) | 19.81 | <0.01 | 0.01 |
|  |  |  |  | SDQ (Anxiety) | 3368 | 1778 | | | 1590 | 3.30 (2.37) | 3.46 (2.41) | 3.13 (2.32) | 15.50 | <0.01 | 0.00 |
|  |  |  |  | CAST (Social) | 3249 | 1726 | | | 1523 | 4.38 (2.31) | 4.06 (2.25) | 4.74 (2.32) | 71.65 | <0.01 | 0.02 |
|  |  |  |  | CAST (Non-Social) | 3260 | 1734 | | | 1526 | 3.92 (2.10) | 3.83 (2.06) | 4.02 (2.14) | 6.59 | 0.01 | 0.00 |
|  |  |  |  | CAST (Communication) | 3255 | 1732 | | | 1523 | 3.43 (2.55) | 3.21 (2.50) | 3.68 (2.59) | 28.81 | <0.01 | 0.01 |
| **12** | | Parent | P factor | SDQ (Prosocial) | 5865 | 3081 | | | 2784 | 8.54 (1.65) | 8.78 (1.53) | 8.26 (1.73) | 146.83 | <0.01 | 0.02 |
|  |  |  |  | SDQ (Hyperactivity) | 5850 | 3072 | | | 2778 | 2.86 (2.29) | 2.37 (2.03) | 3.41 (2.42) | 314.83 | <0.01 | 0.05 |
|  |  |  |  | SDQ (Conduct) | 5850 | 3072 | | | 2778 | 1.34 (1.49) | 1.24 (1.40) | 1.46 (1.57) | 32.96 | <0.01 | 0.01 |
|  |  |  |  | SDQ (Peer Problems) | 5849 | 3072 | | | 2777 | 1.12 (1.55) | 1.00 (1.43) | 1.25 (1.66) | 37.34 | <0.01 | 0.01 |
|  |  |  |  | SDQ (Anxiety) | 5850 | 3072 | | | 2778 | 1.85 (1.95) | 1.98 (2.01) | 1.70 (1.87) | 29.24 | <0.01 | 0.00 |
|  |  |  |  | MFQ | 5848 | 3072 | | | 2776 | 1.18 (2.31) | 1.22 (2.39) | 1.14 (2.21) | 2.06 | 0.15 | 0.00 |
|  |  |  |  | APSD (Callous-Unemotional) | 5864 | 3081 | | | 2783 | 3.09 (1.93) | 2.90 (1.86) | 3.29 (1.98) | 62.02 | <0.01 | 0.01 |
|  |  |  |  | APSD (Narcissism) | 5863 | 3080 | | | 2783 | 1.51 (1.78) | 1.32 (1.63) | 1.72 (1.92) | 74.78 | <0.01 | 0.01 |
|  |  |  |  | APSD (Impulsivity) | 5847 | 3073 | | | 2774 | 2.57 (1.90) | 2.25 (1.78) | 2.93 (1.97) | 195.06 | <0.01 | 0.03 |
|  |  |  |  | CAST (Social) | 6211 | 3224 | | | 2987 | 1.64 (1.55) | 1.32 (1.36) | 1.99 (1.66) | 301.93 | <0.01 | 0.05 |
|  |  |  |  | CAST (Non-Social) | 6203 | 3223 | | | 2980 | 1.43 (1.32) | 1.31 (1.25) | 1.56 (1.38) | 59.88 | <0.01 | 0.01 |
|  |  |  |  | CAST (Communication) | 6212 | 3225 | | | 2987 | 1.96 (1.92) | 1.84 (1.79) | 2.08 (2.04) | 24.92 | <0.01 | 0.00 |
|  |  |  |  | CBRS (Hyperactivity-Impulsivity) | 5855 | 3078 | | | 2777 | 4.34 (4.49) | 3.66 (3.83) | 5.10 (5.02) | 152.99 | <0.01 | 0.03 |
|  |  |  |  | CBRS (Inattention) | 5859 | 3081 | | | 2778 | 5.63 (5.21) | 4.57 (4.59) | 6.81 (5.59) | 281.58 | <0.01 | 0.05 |
|  |  | Twin | Home environment | Parental discipline | 5863 | 3078 | | | 2785 | 3.13 (1.48) | 3.04 (1.45) | 3.23 (1.50) | 22.73 | <0.01 | 0.00 |
|  |  |  |  | CHAOS | 5867 | 3079 | | | 2788 | 4.01 (2.06) | 3.87 (2.06) | 4.16 (2.04) | 28.90 | <0.01 | 0.00 |
|  |  |  | P factor | SDQ (Prosocial) | 5841 | 3059 | | | 2782 | 7.45 (1.92) | 7.90 (1.74) | 6.95 (1.98) | 381.79 | <0.01 | 0.06 |
|  |  |  |  | SDQ (Hyperactivity) | 5839 | 3059 | | | 2780 | 3.55 (2.31) | 3.14 (2.14) | 4.01 (2.41) | 209.47 | <0.01 | 0.03 |
|  |  |  |  | SDQ (Conduct) | 5839 | 3059 | | | 2780 | 1.92 (1.66) | 1.68 (1.52) | 2.18 (1.77) | 135.82 | <0.01 | 0.02 |
|  |  |  |  | SDQ (Peer Problems) | 5840 | 3059 | | | 2781 | 1.37 (1.60) | 1.28 (1.57) | 1.47 (1.62) | 21.96 | <0.01 | 0.00 |
|  |  |  |  | SDQ (Anxiety) | 5837 | 3058 | | | 2779 | 2.22 (2.07) | 2.41 (2.13) | 2.02 (1.99) | 51.66 | <0.01 | 0.01 |
|  |  |  |  | MFQ | 5857 | 3073 | | | 2784 | 2.35 (3.37) | 2.32 (3.47) | 2.38 (3.26) | 0.51 | 0.48 | 0.00 |
| **16** | | Parent | P factor | SDQ (Prosocial) | 5122 | 2818 | | | 2304 | 8.23 (1.95) | 8.51 (1.80) | 7.88 (2.07) | 137.04 | <0.01 | 0.03 |
|  |  |  |  | SDQ (Hyperactivity) | 5116 | 2814 | | | 2302 | 2.29 (1.99) | 1.98 (1.81) | 2.68 (2.13) | 161.72 | <0.01 | 0.03 |
|  |  |  |  | SDQ (Conduct) | 5126 | 2820 | | | 2306 | 1.23 (1.38) | 1.19 (1.35) | 1.28 (1.41) | 5.43 | 0.02 | 0.00 |
|  |  |  |  | ARBQ | 5127 | 2818 | | | 2309 | 3.67 (4.32) | 4.27 (4.63) | 2.94 (3.78) | 123.15 | <0.01 | 0.02 |
|  |  |  |  | MFQ | 5123 | 2817 | | | 2306 | 1.03 (2.37) | 1.22 (2.69) | 0.78 (1.89) | 44.45 | <0.01 | 0.01 |
|  |  |  |  | CBRS (Impulsivity) | 5119 | 2815 | | | 2304 | 2.63 (3.52) | 2.47 (3.26) | 2.83 (3.81) | 13.27 | <0.01 | 0.00 |
|  |  |  |  | CBRS (Inattention) | 5120 | 2815 | | | 2305 | 4.29 (4.99) | 3.40 (4.30) | 5.39 (5.52) | 210.85 | <0.01 | 0.04 |
|  |  |  |  | ICUT (Callous) | 5122 | 2816 | | | 2306 | 4.75 (3.68) | 4.32 (3.46) | 5.27 (3.86) | 86.94 | <0.01 | 0.02 |
|  |  |  |  | ICUT (Unemotional) | 5127 | 2818 | | | 2309 | 5.20 (2.94) | 4.69 (2.79) | 5.84 (2.99) | 202.93 | <0.01 | 0.04 |
|  |  |  |  | ICUT (Uncaring) | 5127 | 2818 | | | 2309 | 7.68 (4.83) | 6.81 (4.57) | 8.74 (4.94) | 212.24 | <0.01 | 0.04 |
|  |  |  |  | AQ (Social) | 5125 | 2817 | | | 2308 | 7.36 (4.76) | 6.93 (4.57) | 7.87 (4.94) | 49.74 | <0.01 | 0.01 |
|  |  |  |  | AQ (Attention Switching) | 5122 | 2815 | | | 2307 | 8.31 (4.28) | 7.84 (4.09) | 8.88 (4.43) | 76.55 | <0.01 | 0.01 |
|  |  |  |  | AQ (Imagination) | 5101 | 2810 | | | 2291 | 4.39 (3.46) | 3.85 (3.20) | 5.05 (3.64) | 157.02 | <0.01 | 0.03 |
|  |  |  |  | AQ (Attention to Detail) | 5085 | 2810 | | | 2275 | 4.89 (3.54) | 4.51 (3.42) | 5.36 (3.63) | 71.98 | <0.01 | 0.01 |
|  |  | Twin | Home environment | Parental discipline | 2745 | 1023 | | | 1452 | 3.09 (1.33) | 3.06 (1.31) | 3.11 (1.35) | 1.02 | 0.31 | 0.00 |
|  |  |  |  | CHAOS | 2782 | 1602 | | | 1180 | 4.13 (2.04) | 4.15 (2.06) | 4.12 (2.02) | 0.11 | 0.74 | 0.00 |
|  |  |  | P factor | SDQ (Prosocial) | 5088 | 2808 | | | 2280 | 7.13 (1.95) | 7.61 (1.80) | 6.53 (1.96) | 415.23 | <0.01 | 0.08 |
|  |  |  |  | SDQ (Hyperactivity) | 5089 | 2808 | | | 2281 | 3.59 (2.31) | 3.57 (2.33) | 3.62 (2.30) | 0.67 | 0.41 | 0.00 |
|  |  |  |  | SDQ (Conduct) | 5089 | 2808 | | | 2281 | 1.65 (1.46) | 1.56 (1.43) | 1.77 (1.50) | 25.24 | <0.01 | 0.00 |
|  |  |  |  | SDQ (Peer Problems) | 5091 | 2809 | | | 2282 | 1.57 (1.52) | 1.52 (1.50) | 1.63 (1.54) | 6.60 | 0.01 | 0.00 |
|  |  |  |  | SDQ (Anxiety) | 5090 | 2808 | | | 2282 | 2.76 (2.26) | 3.41 (2.33) | 1.96 (1.88) | 582.80 | <0.01 | 0.10 |
|  |  |  |  | MFQ | 5097 | 2815 | | | 2282 | 3.69 (4.54) | 4.50 (5.05) | 2.70 (3.56) | 205.38 | <0.01 | 0.04 |
|  |  |  |  | AQ (Social) | 5087 | 2810 | | | 2277 | 7.05 (4.21) | 7.11 (4.36) | 6.97 (4.01) | 1.41 | 0.24 | 0.00 |
|  |  |  |  | AQ (Attention to Detail) | 5085 | 2810 | | | 2275 | 4.89 (3.54) | 4.51 (3.42) | 5.36 (3.63) | 71.98 | <0.01 | 0.01 |
|  | Note: *n* = number of participants, SD = standard deviation, Group difference= F statistic | | | | | | | | | | | | | | |

#### Table S2. *Factor loadings for the p factor model across ages and raters.*

| **Model** | **Measure** | **Estimate** | **z-value** | **P (>\|z\|)** |
| --- | --- | --- | --- | --- |
| Parent p at 9 | SDQ (Hyperactivity) | 0.644 | 39.505 | ≤ 0.01 |
|  | SDQ (Conduct) | 0.726 | 46.449 | ≤ 0.01 |
|  | SDQ (Peer problems) | 0.479 | 27.640 | ≤ 0.01 |
|  | SDQ (emotional problems) | 0.445 | 25.594 | ≤ 0.01 |
|  | APSD (callous-unemotional) | 0.411 | 23.388 | ≤ 0.01 |
|  | APSD (narcissism) | 0.709 | 44.923 | ≤ 0.01 |
|  | APSD (impulsivity) | 0.759 | 49.466 | ≤ 0.01 |
|  | RPAQ (proactive aggression) | 0.570 | 34.062 | ≤ 0.01 |
|  | RPAQ (reactive aggression) | 0.635 | 38.885 | ≤ 0.01 |
|  | CAST (social) | 0.348 | 19.442 | ≤ 0.01 |
|  | CAST (non-social) | 0.319 | 17.782 | ≤ 0.01 |
|  | CAST (communication) | 0.602 | 36.391 | ≤ 0.01 |
|  | SDQ Prosocial | -0.345 | -19.344 | ≤ 0.01 |
| Child p at 9 | SDQ (Hyperactivity) | 0.525 | 28.126 | ≤ 0.01 |
|  | SDQ (Conduct) | 0.588 | 32.001 | ≤ 0.01 |
|  | SDQ (Peer problems) | 0.541 | 29.381 | ≤ 0.01 |
|  | SDQ (emotional problems) | 0.609 | 33.963 | ≤ 0.01 |
|  | CAST (social) | 0.266 | 13.410 | ≤ 0.01 |
|  | CAST (non-social) | 0.538 | 28.372 | ≤ 0.01 |
|  | CAST (communication) | 0.721 | 40.168 | ≤ 0.01 |
|  | SDQ Prosocial | -0.169 | -8.422 | ≤ 0.01 |
| Parent p at 12 | SDQ (Hyperactivity) | 0.777 | 67.169 | ≤ 0.01 |
|  | SDQ (Conduct) | 0.647 | 52.363 | ≤ 0.01 |
|  | SDQ (Peer Problems) | 0.469 | 35.611 | ≤ 0.01 |
|  | SDQ (Emotional Problems) | 0.426 | 32.039 | ≤ 0.01 |
|  | MFQ | 0.532 | 41.250 | ≤ 0.01 |
|  | APSD (Callous-Unemotional) | 0.418 | 31.467 | ≤ 0.01 |
|  | APSD (Narcissism) | 0.575 | 45.131 | ≤ 0.01 |
|  | APSD (Impulsivity) | 0.769 | 66.267 | ≤ 0.01 |
|  | CAST (Social) | 0.262 | 19.490 | ≤ 0.01 |
|  | CAST (Non-Social) | 0.350 | 26.433 | ≤ 0.01 |
|  | CAST (Communication) | 0.617 | 50.730 | ≤ 0.01 |
|  | Conners (Hyperactivity) | 0.708 | 58.992 | ≤ 0.01 |
|  | Conners (Inattention) | 0.734 | 61.814 | ≤ 0.01 |
|  | SDQ Prosocial | -0.372 | -27.642 | ≤ 0.01 |
| Child p at 12 | SDQ (Hyperactivity) | 0.559 | 38.994 | ≤ 0.01 |
|  | SDQ (Conduct) | 0.601 | 42.149 | ≤ 0.01 |
|  | SDQ (Peer Problems) | 0.528 | 38.043 | ≤ 0.01 |
|  | SDQ (Emotional Problems) | 0.651 | 48.062 | ≤ 0.01 |
|  | MFQ | 0.753 | 56.459 | ≤ 0.01 |
|  | SDQ Prosocial | -0.213 | -14.054 | ≤ 0.01 |
| Parent p at 16 | SDQ (Hyperactivity) | 0.719 | 56.015 | ≤ 0.01 |
|  | SDQ (Conduct) | 0.640 | 48.371 | ≤ 0.01 |
|  | ARBQ Anxiety | 0.480 | 33.789 | ≤ 0.01 |
|  | MFQ | 0.502 | 35.897 | ≤ 0.01 |
|  | Conners hyperactivity | 0.485 | 34.360 | ≤ 0.01 |
|  | Conners inattention | 0.728 | 56.857 | ≤ 0.01 |
|  | ICUT (Callous) | 0.593 | 43.659 | ≤ 0.01 |
|  | ICUT (Unemotional) | 0.392 | 27.077 | ≤ 0.01 |
|  | ICUT (Uncaring) | 0.715 | 55.258 | ≤ 0.01 |
|  | AQ (Social) | 0.441 | 30.594 | ≤ 0.01 |
|  | AQ (Attention Switching) | 0.576 | 41.908 | ≤ 0.01 |
|  | AQ (Imagination) | 0.477 | 33.919 | ≤ 0.01 |
|  | AQ (Attention to Detail) | 0.011 | 0.700 | 0.484 |
|  | SDQ Prosocial | -0.595 | -43.596 | ≤ 0.01 |
| Child p at 16 | SDQ (Hyperactivity) | 0.453 | 30.149 | ≤ 0.01 |
|  | SDQ (Conduct) | 0.402 | 26.315 | ≤ 0.01 |
|  | SDQ (Peer Problems) | 0.527 | 35.061 | ≤ 0.01 |
|  | SDQ (Emotional Problems) | 0.762 | 56.379 | ≤ 0.01 |
|  | MFQ | 0.773 | 56.923 | ≤ 0.01 |
|  | AQ (Social) | 0.500 | 32.637 | ≤ 0.01 |
|  | AQ (Attention to Detail) | 0.206 | 13.250 | ≤ 0.01 |
|  | SDQ Prosocial | -0.214 | -13.410 | ≤ 0.01 |

#### **Table S3.** *Model fit statistics for the p factor model across ages and raters.*

| **Model fit** | **Parent p at 9** | **Child p at 9** | **Parent p at 12** | **Child p at 12** | **Parent p at 16** | **Child p at 16** |
| --- | --- | --- | --- | --- | --- | --- |
| AIC | 115000.041 | 71502.478 | 213828.906 | 93213.877 | 186465.404 | 109163.31 |
| BIC | 115239.308 | 71649.564 | 214111.724 | 93334.065 | 186740.253 | 109320.207 |
| CFI | 0.75 | 0.756 | 0.797 | 0.823 | 0.687 | 0.172 |
| RMSA | 0.127 | 0.138 | 0.108 | 0.16 | 0.139 | 0.09 |
| SRMR | 0.08 | 0.078 | 0.068 | 0.068 | 0.087 | 0.067 |

#### **Table S****4.** Model fit statistics for phenotypic and genetic cross-lagged panel models

| **model** | **ep** | **minus2LL** | **df** | **AIC** | **BIC** | **CFI** | **TLI** | **RMSEA** | **diffLL** | **diffdf** | **p** |
| --- | --- | --- | --- | --- | --- | --- | --- | --- | --- | --- | --- |
| Child p -phenotypic | 23 | 64340.52 | 25723 | 12894.52 | -170638.3 | 0.993 | 0.975 | 0.027 | 31.780 | 4 | <0.001 |
| Parent p -phenotypic | 23 | 63427.84 | 26121 | 11185.84 | -175186.7 | 0.991 | 0.967 | 0.039 | 59.320 | 4 | <0.001 |
| Child p -genetic | 57 | 122243.9 | 51205 | 19833.89 | -344705.3 | 0.997 | 0.997 | 0.006 | 163.448 | 123 | 0.009 |
| Parent p -genetic | 57 | 113862.6 | 51973 | 9916.555 | -360090.2 | 0.997 | 0.997 | 0.008 | 194.408 | 123 | <0.001 |

Note: minus2LL = Minus 2*log-likelihood of the comparison model, df = degrees of freedom, AIC = Akaike information criterion, BIC = Bayesian information criterion.

#### **Table S5**. Twin model-fitting results for univariate analyses parent-rated p factor and home environment. A = additive genetic, C = shared environmental, E = non-shared environmental proportions of the variance.

| **Age** | **Composite** | **A** | **95% CI** | | **C** | **95% CI** | | **E** | **95% CI** | |
| --- | --- | --- | --- | --- | --- | --- | --- | --- | --- | --- |
|  |  |  | **Lower** | **Upper** |  | **Lower** | **Upper** |  | **Lower** | **Upper** |
| 9 | Parent p | 0.51 | 0.47 | 0.58 | 0.36 | 0.30 | 0.40 | 0.13 | 0.11 | 0.14 |
|  | Home environment | 0.19 | 0.13 | 0.27 | 0.41 | 0.35 | 0.46 | 0.39 | 0.36 | 0.42 |
| 12 | Parent p | 0.67 | 0.63 | 0.71 | 0.18 | 0.15 | 0.23 | 0.14 | 0.13 | 0.16 |
|  | Home environment | 0.19 | 0.12 | 0.24 | 0.42 | 0.38 | 0.47 | 0.39 | 0.36 | 0.41 |
| 16 | Parent p | 0.59 | 0.55 | 0.64 | 0.28 | 0.24 | 0.32 | 0.13 | 0.11 | 0.14 |
|  | Home environment | 0.31 | 0.21 | 0.41 | 0.26 | 0.18 | 0.33 | 0.44 | 0.40 | 0.48 |

#### **Table S6.** Percentages of genetic and environmental variance unique to each construct in parent-rated model after accounting for variance shared with previous time points. A = additive genetic, C = shared environmental, E = non-shared environmental proportions of the variance.

| **Age** | **Composite** | **A(%)_spec** | **95% CI** | | **C(%)_spec** | **95% CI** | | **E(%)_spec** | **95% CI** | |
| --- | --- | --- | --- | --- | --- | --- | --- | --- | --- | --- |
|  |  |  | **Lower** | **Upper** |  | **Lower** | **Upper** |  | **Lower** | **Upper** |
| 9 | Parent p | 0.51 | 0.47 | 0.58 | 0.36 | 0.30 | 0.40 | 0.13 | 0.11 | 0.14 |
|  | Home environment | 0.19 | 0.13 | 0.27 | 0.41 | 0.35 | 0.46 | 0.39 | 0.36 | 0.42 |
| 12 | Parent p | 0.27 | 0.23 | 0.32 | 0.03 | 0.00 | 0.06 | 0.12 | 0.11 | 0.13 |
|  | Home environment | 0.12 | 0.02 | 0.20 | 0.19 | 0.12 | 0.24 | 0.39 | 0.36 | 0.41 |
| 16 | Parent p | 0.31 | 0.27 | 0.35 | 0.12 | 0.09 | 0.15 | 0.11 | 0.10 | 0.12 |
|  | Home environment | 0.26 | 0.17 | 0.35 | 0.09 | 0.02 | 0.18 | 0.43 | 0.39 | 0.46 |

#### **Table S7.** Twin model-fitting results for univariate analyses of child-rated p factor and home environment. A = additive genetic, C = shared environmental, E = non-shared environmental proportions of the variance.

| **Age** | **Composite** | **A** | **95% CI** | | **C** | **95% CI** | | **E** | **95% CI** | |
| --- | --- | --- | --- | --- | --- | --- | --- | --- | --- | --- |
|  |  |  | **Lower** | **Upper** |  | **Lower** | **Upper** |  | **Lower** | **Upper** |
| 9 | Child p | 0.47 | 0.41 | 0.51 | 0.15 | 0.11 | 0.19 | 0.39 | 0.36 | 0.41 |
|  | Home environment | 0.20 | 0.13 | 0.28 | 0.41 | 0.35 | 0.46 | 0.39 | 0.36 | 0.42 |
| 12 | Child p | 0.42 | 0.37 | 0.47 | 0.15 | 0.11 | 0.19 | 0.42 | 0.40 | 0.44 |
|  | Home environment | 0.20 | 0.15 | 0.26 | 0.42 | 0.37 | 0.45 | 0.39 | 0.36 | 0.40 |
| 16 | Child p | 0.46 | 0.41 | 0.51 | 0.06 | 0.03 | 0.10 | 0.48 | 0.45 | 0.50 |
|  | Home environment | 0.29 | 0.19 | 0.39 | 0.27 | 0.19 | 0.35 | 0.44 | 0.40 | 0.48 |

#### **Table S8.** Percentages of genetic and environmental variance unique to each construct in child-rated model after accounting for variance shared with previous time points. A = additive genetic, C = shared environmental, E = non-shared environmental proportions of the variance.

| **Age** | **Composite** | **A(%)_spec** | **95% CI** | | **C(%)_spec** | **95% CI** | | **E(%)_spec** | **95% CI** | |
| --- | --- | --- | --- | --- | --- | --- | --- | --- | --- | --- |
|  |  |  | **Lower** | **Upper** |  | **Lower** | **Upper** |  | **Lower** | **Upper** |
| 9 | Child p | 0.47 | 0.41 | 0.51 | 0.15 | 0.11 | 0.19 | 0.39 | 0.36 | 0.41 |
|  | Home environment | 0.20 | 0.13 | 0.28 | 0.41 | 0.35 | 0.46 | 0.39 | 0.36 | 0.42 |
| 12 | Child p | 0.21 | 0.15 | 0.27 | 0.07 | 0.03 | 0.11 | 0.42 | 0.39 | 0.44 |
|  | Home environment | 0.15 | 0.09 | 0.22 | 0.12 | 0.04 | 0.18 | 0.38 | 0.36 | 0.40 |
| 16 | Child p | 0.30 | 0.25 | 0.35 | 0.03 | 0.01 | 0.07 | 0.47 | 0.44 | 0.49 |
|  | Home environment | 0.24 | 0.15 | 0.33 | 0.10 | 0.03 | 0.17 | 0.43 | 0.39 | 0.47 |

### **Supplementary Figures**

#### **Figure S1.** *A correlation heatmap for all phenotypic variables at age 9*

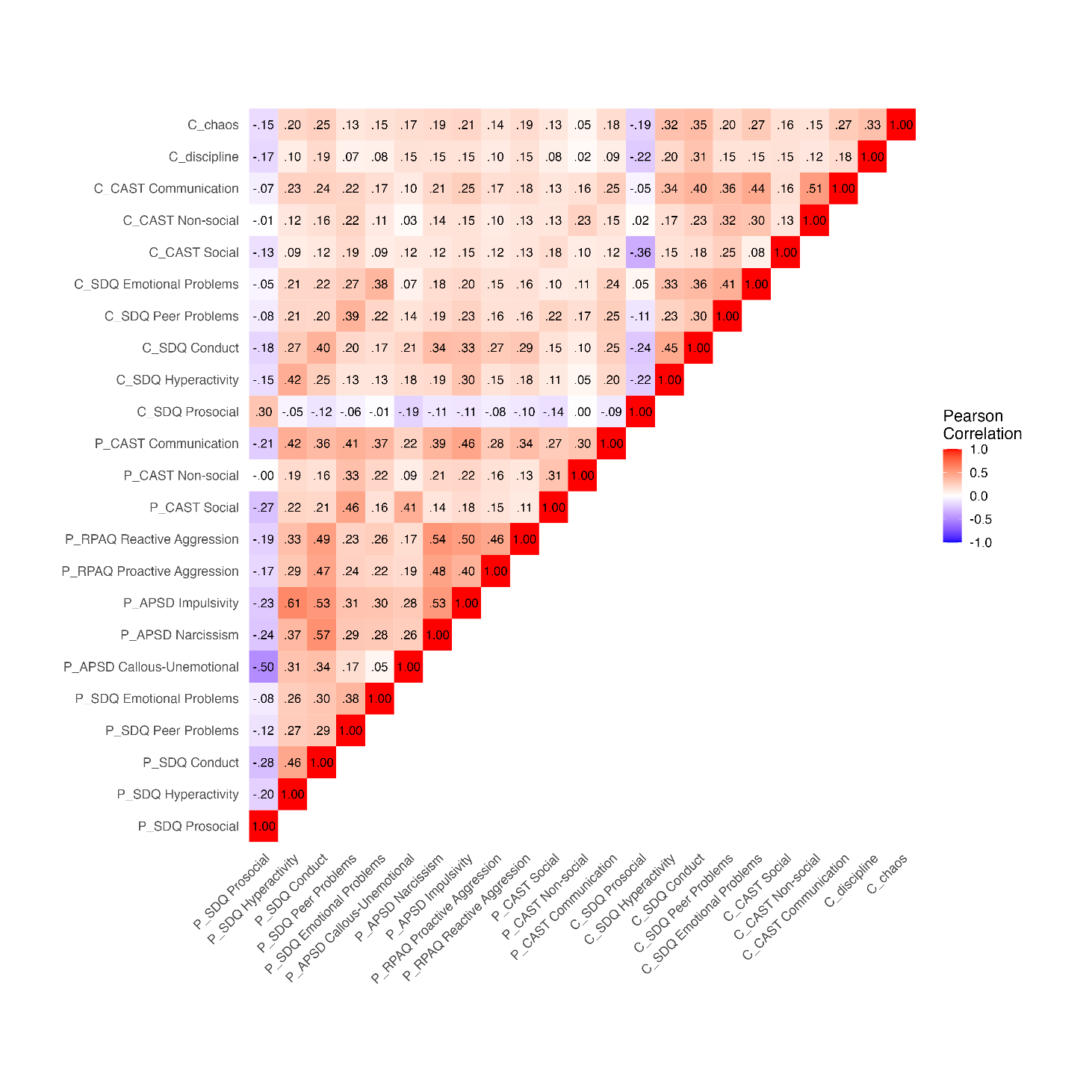

Note: Variable name preceded by P = parent report, C = child-report. CAST = Childhood Autism Spectrum Test, SDQ = Strengths and Difficulties Questionnaire, RPAQ = Reactive-Proactive Aggression Questionnaire, APSD = Anti-Social Process Screening Device.

#### **Figure S2.** *A correlation heatmap for all phenotypic variables at age 12*

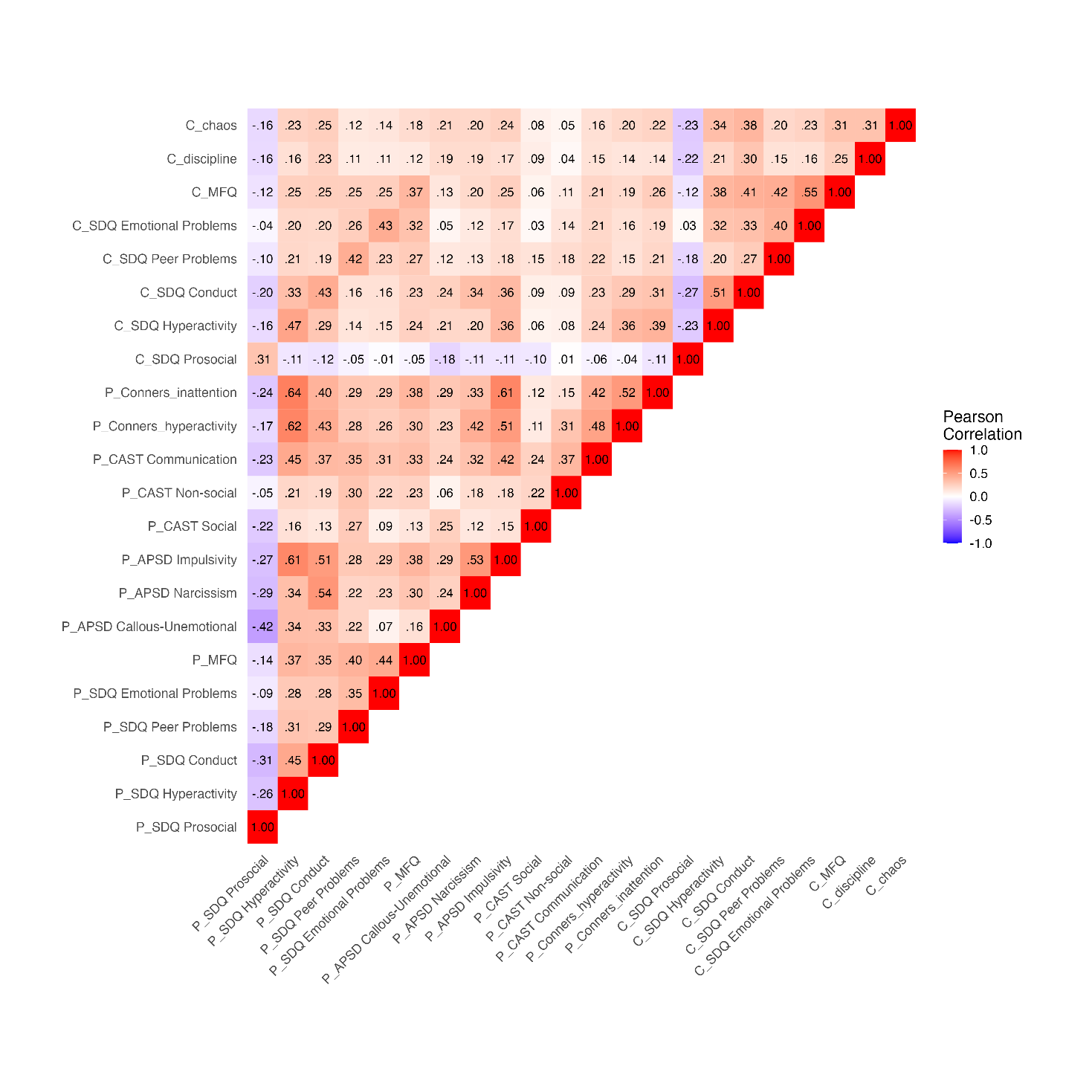

Note: Variable name preceded by P = parent report, C = child-report. SDQ = Strengths and Difficulties Questionnaire, CAST = Childhood Autism Spectrum Test, APSD = Anti-Social Process Screening Device, MFQ = Mood and Feelings Questionnaire.

#### **Figure S3.** A correlation heatmap for all phenotypic variables at age 16

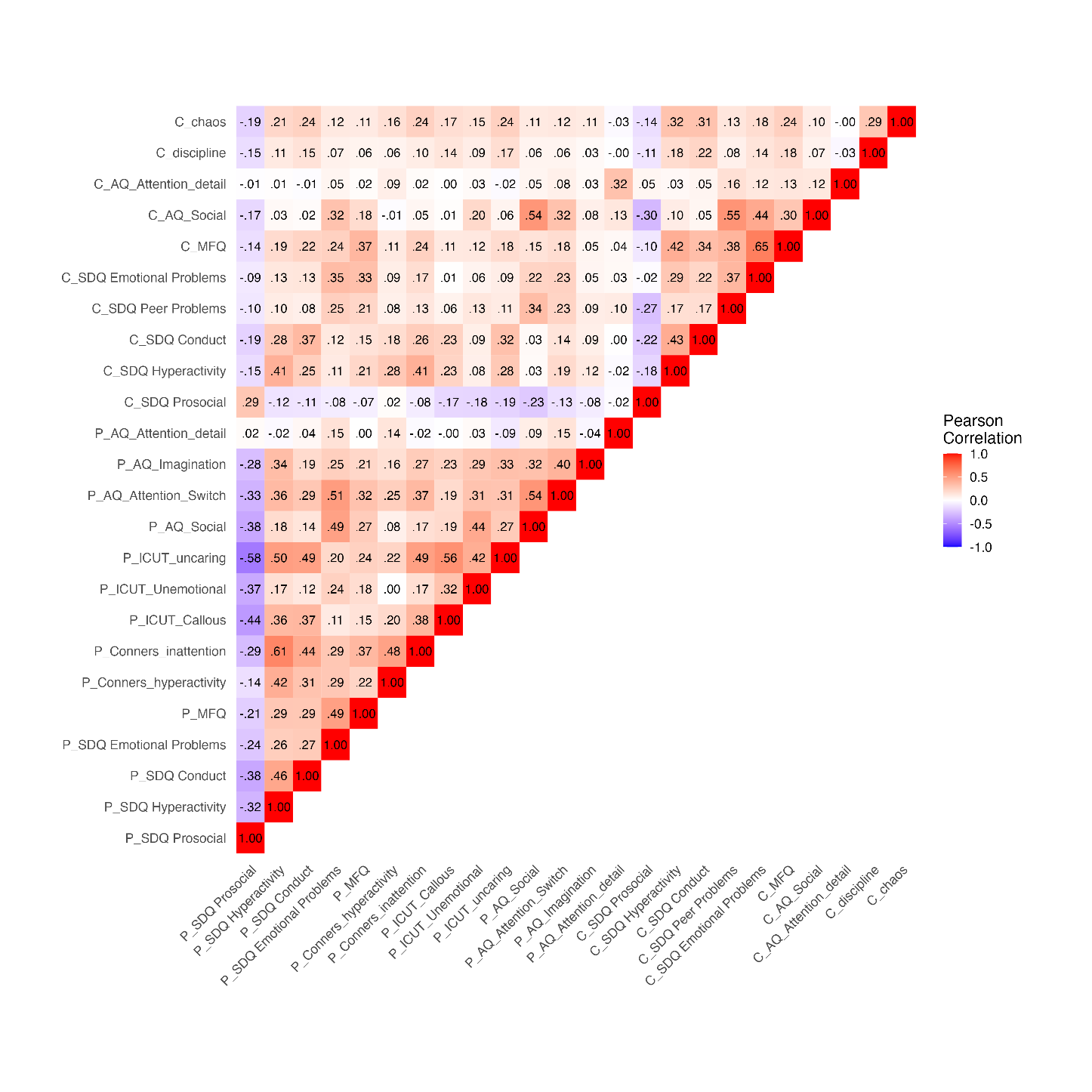

Note: Variable name preceded by P = parent report, C = child-report. AQ = Autism Quotient, MFQ = Mood and Feelings Questionnaire, SDQ = Strengths and Difficulties Questionnaire, ICUT = Inventory of Callous-Unemotional Traits.

#### **Figure S4.** *A correlation heatmap for all composites*

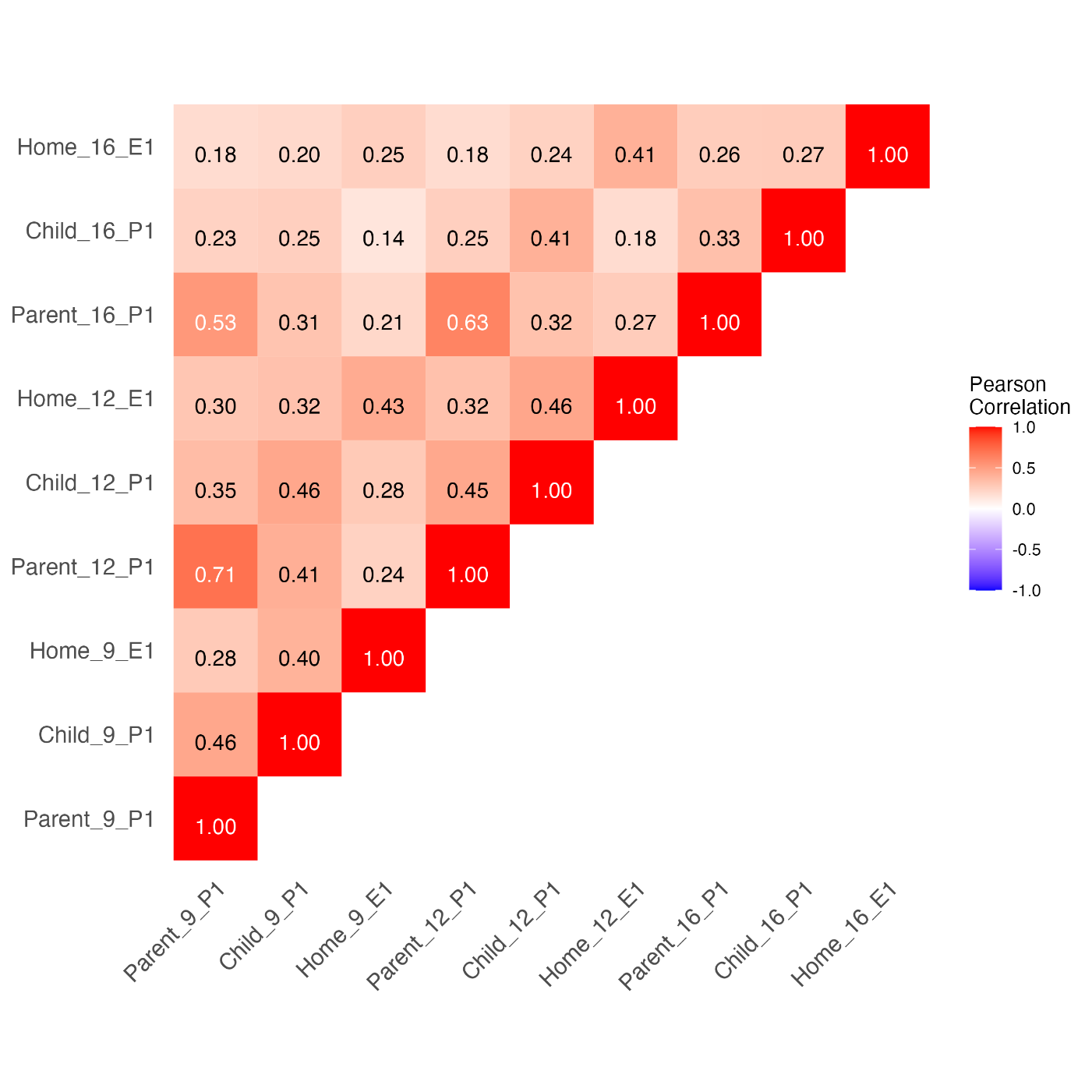

Note: ‘Parent’ and ‘Child’ indicate informant who completed constituent measures. Numbers 9, 12 and 16 denote measurement age. P1 = p factor (1^st^ principal component), E1 = home environment (1^st^ principal component).

#### **Figure S5.** *A cross-lagged panel model between the parent-rated p factor (p) and home environment composite (he) MZ difference scores (Model 3)*

0.22

(0.15-0.28)

-0.02

(-0.08-0.04)

0.06

(0.00-0.12)

0.02

(-0.05-0.08)

0.24

(0.19-0.29)

0.02

(-0.03-0.08)

0.03

(-0.04-0.11)

0.10

(0.03-0.18)

1.00

(1.00-1.00)

1.00

(1.00-1.00)

0.04

(-0.02-0.10)

0.95

(0.92-0.97)

0.94

(0.91-0.96)

1.00

(0.99-1.00)

0.99

(0.97-1.00)

0.06

(0.01-0.10)

0.13

(0.05-0.22)

Note: 95% CI are in brackets. Solid lines indicate statistical significance; dashed lines indicate non-significance paths estimates

#### **Figure S6.** *A cross-lagged panel model between the twin-rated p factor (p) and home environment composite (he) MZ difference scores (Model 4)*

1.00

(1.00-1.00)

1.00

(1.00-1.00)

0.10

(0.03-0.17)

0.09

(0.02-0.15)

0.02

(-0.05-0.08)

0.00

(-0.06-0.06)

0.09

(0.04-0.14)

0.04

(-0.04-0.13)

0.02

(-0.03-0.07)

0.09

(0.02-0.17)

0.23

(0.17-0.29)

0.99

(0.97-1.00)

0.99

(0.98-1.00)

0.99

(0.97-1.00)

0.99

(0.98-1.00)

0.08

(0.01-0.15)

0.20

(0.15-0.24)

Note: 95% CI are in brackets. Solid lines indicate statistical significance where 95% confidence intervals did not cross zero; dashed lines indicate non-significance as a result of crossing zero.

### **Procedure**

To capture the p factor, the following constructs were used: *ADHD* (Conners’ Comprehensive Behavioural Rating Scale [CBRS]; Conners, 1970), *aggression* (Reactive-Proactive Aggression Questionnaire [RPAQ]; Raine et al., 2006),; Eley et al., 2003), *autism traits* (Childhood Autism Spectrum Test [CAST]; Scott, Baron-Cohen, Bolton & Brayne, 2002; Autism Quotient [AQ]; Baron-Cohen, Wheelwright, Skinner, Martin & Clubley, 2001), *callous-unemotional traits* (Inventory for the Callous Unemotional Scale [ICUT]; Frick, 2004), *depression* (Moods and Feelings Questionnaire [MFQ]; Angold, Costello, Pickles & Winder, 1987)*, emotional problems* (Strengths and Difficulties Questionnaire [SDQ – Anxiety subscale]; Goodman, 1997)*, hyperactivity* (SDQ – hyperactivity subscale)*, peer problems* (SDQ – peer problems subscale)*, prosocial behaviour* (SDQ – prosocial subscale, reversed) and *psychopathic tendencies* (Anti-Social Process Screening Device [APSD]; Frick & Hare, 2001). All measures are well-validated and have previously demonstrated sound psychometric properties, with moderate to high levels of internal consistency (all Cronbach’s alpha > 0.6). Different measures were used at different waves to capture parallel constructs at each wave whilst maintaining developmental appropriateness.

To capture home environment, a composite was calculated using the following constructs at all three waves: *household chaos* (Confusion, Hubbub and Order Scale [CHAOS]; Matheny Jr et al., 1995) and *parental discipline* (derived from Deater-Deckard, Dodge, Bates & Pettitt, 1998). These measures were chosen as they have been consistently administered at each selected wave. A summary of the measures administered at each wave can be found in Table 1P.

| **Table 1P.** *Summary of measures included to calculate p-factor and home environment composites* | | | |
| --- | --- | --- | --- |
| **Wave age** | **p factor measures** |  | **Home environment measures (all twin-rated)** |
|  | **Parent-rated** | **Twin-rated** |  |
| 9 | SDQ Emotional Problems  SDQ Prosocial  SDQ Hyperactivity  SDQ Conduct  SDQ Peer Problems  CAST Social  CAST Non-Social  CAST Communication  APSD Impulsivity  APSD Narcissism  RPAQ Reactive Aggression  RPAQ Proactive Aggression | SDQ Emotional Problems  SDQ Prosocial  SDQ Hyperactivity  SDQ Conduct  SDQ Peer Problems  CAST Social  CAST Non-Social  CAST Communication | CHAOS  Parental discipline |
| 12 | SDQ Emotional Problems  SDQ Prosocial  SDQ Hyperactivity  SDQ Conduct  SDQ Peer Problems  MFQ  CAST Social  CAST Non-Social  CAST Communication  APSD Impulsivity  APSD Callous-Unemotional  APSD Narcissism  CBRS Inattention  CBRS Hyperactivity | SDQ Emotional Problems  SDQ Prosocial  SDQ Hyperactivity  SDQ Conduct  SDQ Peer Problems  MFQ | CHAOS  Parental discipline |
| 16 | SDQ Emotional Problems SDQ Prosocial  SDQ Hyperactivity  SDQ Conduct  AQ Attention to Detail  AQ Imagination  AQ Attention Switching  AQ Social  MFQ  CBRS Inattention  CBRS Hyperactivity ICUT Uncaring  ICUT Unemotional  ICUT Callous | SDQ Emotional Problems  SDQ Prosocial  SDQ Hyperactivity  SDQ Conduct  SDQ Peer Problems  MFQ  AQ Attention to Detail  AQ Social | CHAOS  Parental discipline |

Data for each wave in the present study were collected from twins and parents by TEDS via a test booklet, with an additional wellbeing questionnaire administered digitally to twins at 16. The dataset was exported to an SPSS (IBM Statistical Product and Service Solutions 26) file. Measures were adjusted for age and sex, and descriptive statistics were generated. Correlations and cross-lagged panel model analyses were conducted using R version 3.6.3 (R Core Team, 2020a). The following packages were in the main analyses: lavaan (Rosseel, 2012), psych (v2.1.3; Revelle, 2020), ggplot2 (Wickham, 2016). The following packages were used for cross-lagged analyses: devtools (v2.4.0; Wickham, Hester & Chang, 2021), tidyverse (Wickham et al., 2019), OpenMX (v2.0; Neale et al., 2016; Pritikin, Hunter & Boker, 2015; Hunter, 2018), openxlsx (v4.2.3; Schaueberger & Walker, 2020), mlth.data.frame (v1.2; Voronin, 2020), and TwinAnalysis (v0.2.0; Voronin, 2021).

### **References**

Angold, A., Costello, E.J., Pickles, A. & Winder, F. (1987). The development of a questionnaire for use in epidemiological studies of depression in children and adolescents. London: Medical Research Council Child Psychiatry Unit.

Baron-Cohen, S., Wheelwright, S., Skinner, R., Martin, J., & Clubley, E. (2001). The autism-spectrum quotient (AQ): Evidence from Asperger syndrome/high-functioning autism, males and females, scientists and mathematicians. Journal of Autism and Developmental Disorders, 31(1), 5-17.

Conners, C. (1970). Symptom patterns in hyperkinetic, neurotic, and normal children. Child Development, 41, 667-682.

Deater-Deckard, K., Dodge, K. A., Bates, J. E., & Pettit, G. S. (1998). Multiple risk factors in the development of externalising behaviour problems: Group and individual differences. Development and Psychopathology, 10(3), 469 - 493.

Eley, T. C., Bolton, D., O’Connor, T. G., Perrin, S., Smith, P., & Plomin, R. (2003). A twin study of anxiety‐related behaviours in pre‐school children. Journal of Child Psychology and Psychiatry, 44(7), 945-960.

Frick, P. J., & Hare, R. D. (2001). The antisocial process screening device. Toronto: Multi-Health Systems.

Frick, P. J. (2004). The inventory of callous-unemotional traits. Unpublished rating scale.

Goodman, R. (1997). The Strengths and Difficulties Questionnaire: a research note. Journal of Child Psychology And Psychiatry, 38(5), 581-586.

Hunter, M. D. (2018). State space modeling in an open source, modular, structural equation modeling environment. Structural Equation Modeling, 25(2), 307-324.

Matheny Jr, A. P., Wachs, T. D., Ludwig, J. L., & Phillips, K. (1995). Bringing order out of chaos: Psychometric characteristics of the confusion, hubbub, and order scale. Journal of Applied Developmental Psychology, 16(3), 429-444.

Neale, M. C., Hunter, M. D., Pritikin, J. N., Zahery, M., Brick, T. R., Kirkpatrick, R. M., Estabrook, R., Bates, T. C., Maes, H. M., & Boker, S. M. (2016). OpenMx 2.0: Extended structural equation and statistical modeling. Psychometrika, 81(2), 535-549.

Pritikin, J. N., Hunter, M. D., & Boker, S. M. (2015). Modular open-source software for Item Factor Analysis. Educational and Psychological Measurement, 75(3), 458-474.

R Core Team (2020a). R: A language and environment for statistical computing. R Foundation for Statistical Computing, Vienna, Austria. <https://www.R-project.org/>.

R Core Team (2020b). foreign: Read Data Stored by 'Minitab', 'S', 'SAS', 'SPSS', 'Stata', 'Systat', 'Weka', 'dBase', ... R package version 0.8-75. <https://CRAN.R-project.org/package=foreign>

Raine, A., Dodge, K., Loeber, R., Gatzke‐Kopp, L., Lynam, D., Reynolds, C., Stouthamer‐Loeber, M., & Liu, J. (2006). The reactive–proactive aggression questionnaire: Differential correlates of reactive and proactive aggression in adolescent boys. Aggressive Behavior: Official Journal of the Interna-tional Society for Research on Aggression, 32(2), 159-171.

Revelle, W. (2020) psych: Procedures for Personality and Psychological Research. Northwestern University: Evanston, Illinois, USA. <https://CRAN.R-project.org/package=psych>

Rosseel, Y. (2012). lavaan: An R Package for Structural Equation Modeling. Journal of Statistical Software, 48(2), 1-36. <https://www.jstatsoft.org/v48/i02/>

Schauberger, P., & Walker, A. (2020). openxlsx: Read, Write and Edit xlsx Files. R package version 4.2.3. <https://CRAN.R-project.org/package=openxlsx>

Schloerke, B., Cook, D., Larmarange, J., Briatte, F., Marbach, M., Thoen, E., Elberg, A., & Crowley, J. (2021). GGally: Extension to 'ggplot2'. R package ver-sion 2.1.1. <https://CRAN.R-project.org/package=GGally>

Scott, F. J., Baron-Cohen, S., Bolton, P., & Brayne, C. (2002). The CAST (Childhood Asperger Syndrome Test) Preliminary development of a UK screen for mainstream primary-school-age children. Autism, 6(1), 9-31.

Voronin I. (2020). mlth.data.frame: Multi-headed Data Frame. R package version 1.2. Retrieved from: <https://github.com/IvanVoronin/TwinAnalysis>

Voronin, I. (2021). TwinAnalysis: This is a package to simplify structural equation modeling - and particularly, twin analysis - in R. R package version 0.2.0. Retrieved from: <https://github.com/IvanVoronin/TwinAnalysis>

Wickham, H. (2016). ggplot2: Elegant Graphics for Data Analysis. Springer-Verlag: New York.

Wickham, H., Averick, M., Bryan, J., Chang, W., McGowan, L. D. A., François, R., Grolemund, G., Hayes, A., Henry, L., Hester, J., Kuhn, M., Pedersen, T. L., Miller, E., Bache, S. M., Müller, K., Ooms, J., Robinson, D., Seidel, D. P., Spinu, V., Takahashi, K. , Vaughan, D. , Wilke, C., Woo, K., & Yutani, H. (2019). Welcome to the Tidyverse. Journal of Open Source Software, 4(43), 1686. <https://doi.org/10.21105/joss.01686>

Wickham, H., Hester, J., & Chang, W. (2021). devtools: Tools to Make Developing R Packages Easier. R package version 2.4.0. <https://CRAN.R-project.org/package=devtools>
